# Multivariate GWAS reveals shared genetic etiology and pleiotropic loci across carcinomas

**DOI:** 10.1101/2025.09.21.25336284

**Authors:** Yu-Feng Huang, Chenshen Huang

**Author notes:** Correspondence to: Chenshen Huang.

## Abstract

Carcinomas, which arise from epithelial tissues and account for more than 90% of cancers, share molecular programs while exhibiting site-specific biology. However, the genetic partitioning of common versus distinct components remains unclear. We harmonized genome-wide association study (GWAS) summary statistics for 429,158 European-ancestry cases across nine common carcinoma types, and triangulated evidence at the genome-wide, regional, and locus levels to delineate shared and cancer-specific risk. We show that cross-carcinoma overlap is likely systematically underestimated, because loci within the same genomic regions can have discordant effects. To address this, we constructed a cross-carcinoma hierarchical latent-factor model, performed follow-up multivariate GWAS to identify novel pleiotropic loci, and subsequently integrated multi-omics data to prioritize effector genes. This framework partitions general and cancer-specific genetic liability, revealing pleiotropy obscured by conventional analyses. Subsequent multi-omics gene prioritization implicated convergent epithelial growth and differentiation programs, nominating tractable targets for biomarker development, prevention, and mechanism-informed therapies.

## Introduction

Cancer is the second leading cause of death worldwide^1^, with approximately 20 million new cases diagnosed in 2022 alone^2^. Carcinomas, which arise from epithelial cells, account for more than 90% of all malignancies and exhibit both cancer-type-specific biology and shared molecular programs that transcend histological boundaries^2–6^. However, the molecules and variants through which carcinomas share versus maintain distinct etiological drivers remain largely unclear.

Carcinomas show a substantial heritable component^7^, and the development of large, ancestrally diverse genome-wide association studies (GWAS) now enables systematic interrogation of inherited risk^7^. Genome-wide correlation analyses have uncovered modest yet significant genetic sharing between specific carcinoma pairs^8–11^, while cross-cancer polygenic risk scores and meta-analyses reveal widespread pleiotropy, implicating common low-effect variants in addition to rare, high-penetrance mutations^9–12^. Nevertheless, most studies have been limited to pairwise genetic correlation at the whole genome, which obscures locus-specific heterogeneity and can be attenuated when trait effects are directionally mixed^13^. Moreover, traditional cross-cancer meta-GWAS are underpowered to detect pleiotropic loci owing to imbalanced sample sizes, mixtures of cancers with different cells of origin^6,9–11^, and the absence of models that explicitly capture the genetic covariance across traits.

Here, we aggregate GWAS summary statistics from 429,158 cases spanning nine common carcinoma types, which together account for more than 56% of annual cancer diagnoses in Europe (excluding non-melanoma skin cancer)^2^. By triangulating multiple complementary analytic methods, we dissect the shared and cancer-specific architecture of carcinomas at the genome-wide, regional, and individual variant levels. We constructed common-factor and hierarchical-factor models and performed latent factor multivariate GWAS to capture general and cluster-specific genetic effects across carcinomas; collectively, these analyses implicated 170 novel pleiotropic loci and prioritized 167 high-confidence effector genes. Incorporating latent clusters significantly improved power in multi-trait multivariate GWAS of cancers with smaller sample sizes, enabling discovery of 44 additional novel loci, five of which were replicated in an independent cohort. These findings refine our understanding of genetic pleiotropy in carcinogenesis and highlight convergent biological pathways that may inform risk stratification and therapeutic development.

## Results

### GWAS data for nine carcinomas

We assembled the largest, most recent GWAS summary statistics for nine European-ancestry epithelial cancers—lung (LC), esophageal (ESCA), colorectal (CRC), ovarian (OC), endometrial (EC), breast (BRCA), renal cell (RCC), thyroid (THCA), and prostate (PRCA)—applying standard quality control as in prior work (see Methods). Effective sample sizes (Neff) ranged from 26,347 to 270,181, and all traits exhibited SNP-heritability Z-scores > 4 **(Table S1).** An overview is shown in **Fig. 1**

**Fig. 1.**
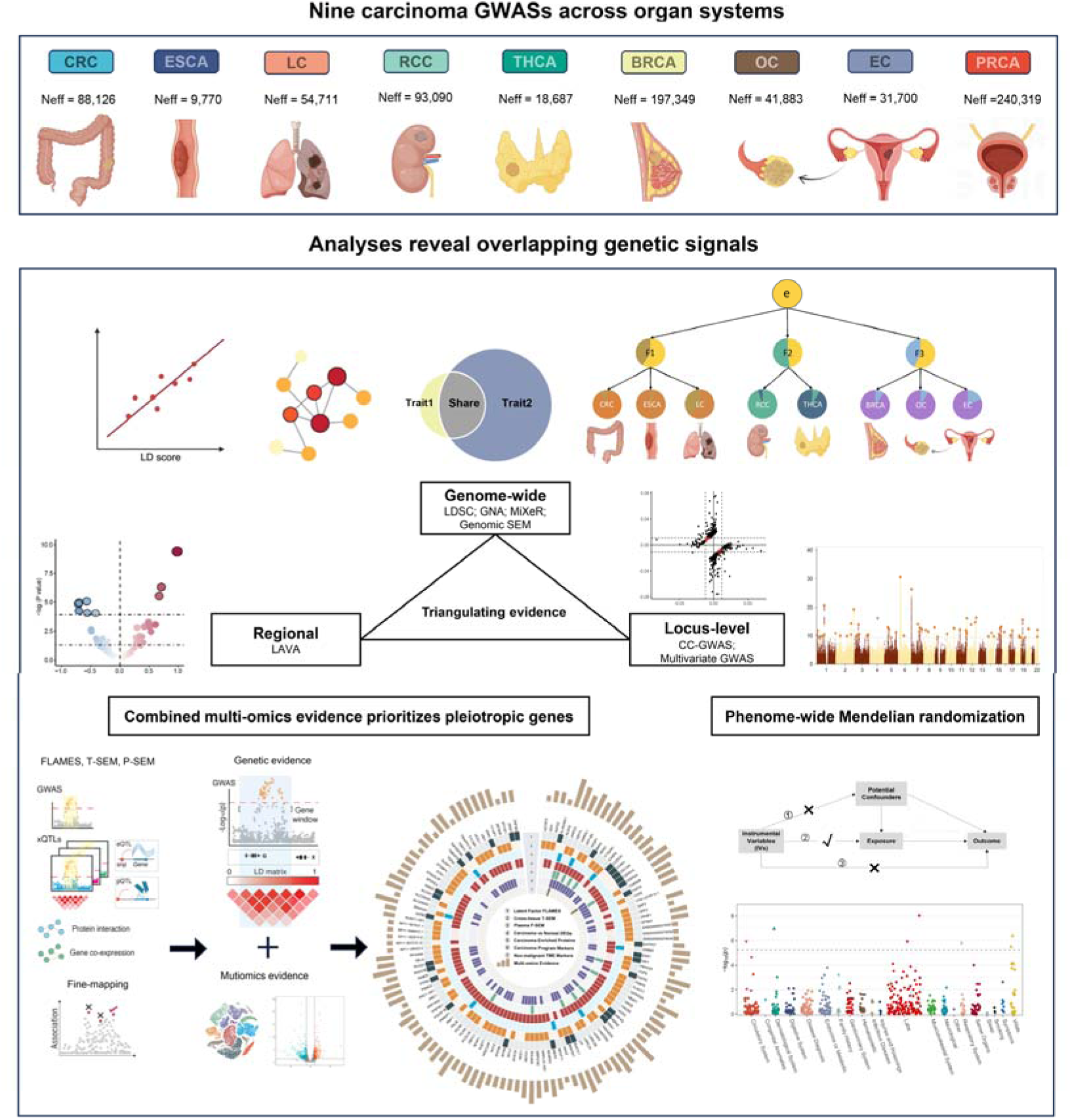
**Study design and flowchart** European-ancestry GWAS datasets for nine carcinomas—breast (BRCA), colorectal (CRC), endometrial (EC), esophageal (ESCA), lung (LC), ovarian (OC), prostate (PRCA), renal cell (RCC), and thyroid (THCA)—were analyzed. LD score regression, LDSC; Genomic Network Analysis, GNA; Fine-mapped Locus Assessment of Multiple Effector genes, FLAMES; case–case GWAS, CC-GWAS; Local Analysis of Variant Association, (LAVA); Genomic Structural Equation Models (GenomicSEM)

### Genome-wide and Conditional Genetic Correlations Among Carcinomas

Using cross-trait LD score regression (LDSC) in nine carcinomas, we estimated pairwise genetic correlations (*r_g_*). Eighteen pairs remained significant after correction for multiple testing (FDR<0.05), with most correlations of modest magnitude (*r_g_* < 0.4), consistent with prior reports^9^ **(Fig. 2a)**. To dissect cancer-pair–specific sharing, we constructed a genetic network of partial correlations. After conditioning on shared variance, eight edges remained significant and were mostly moderate in magnitude; the strongest was OC–EC (estimate = 0.442, SE = 0.106, FDR = 4.21E-8) **(Fig. 2b)**. Notably, once broad correlations were removed, PRCA showed a negative partial correlation with LC (estimate = –0.114, SE = 0.047, FDR = 1.59E-2), indicating that both concordant and discordant effects shape cross-cancer relationships. We next mapped pairwise carcinoma local genetic correlations across the genome. After Bonferroni correction, 36 regions harbored significant local *r_g_*between carcinoma pairs. PRCA showed the largest number of significant local correlations with other carcinomas (n = 23), including negative regional *r_g_* with LC (n = 5) and BRCA (n = 2) **(Table S2; Fig. 2c)**. These findings imply that genome-wide *r_g_* can understate shared architecture when effects are directionally mixed and highlight cancer-pair–specific regional structure.

**Fig. 2.**
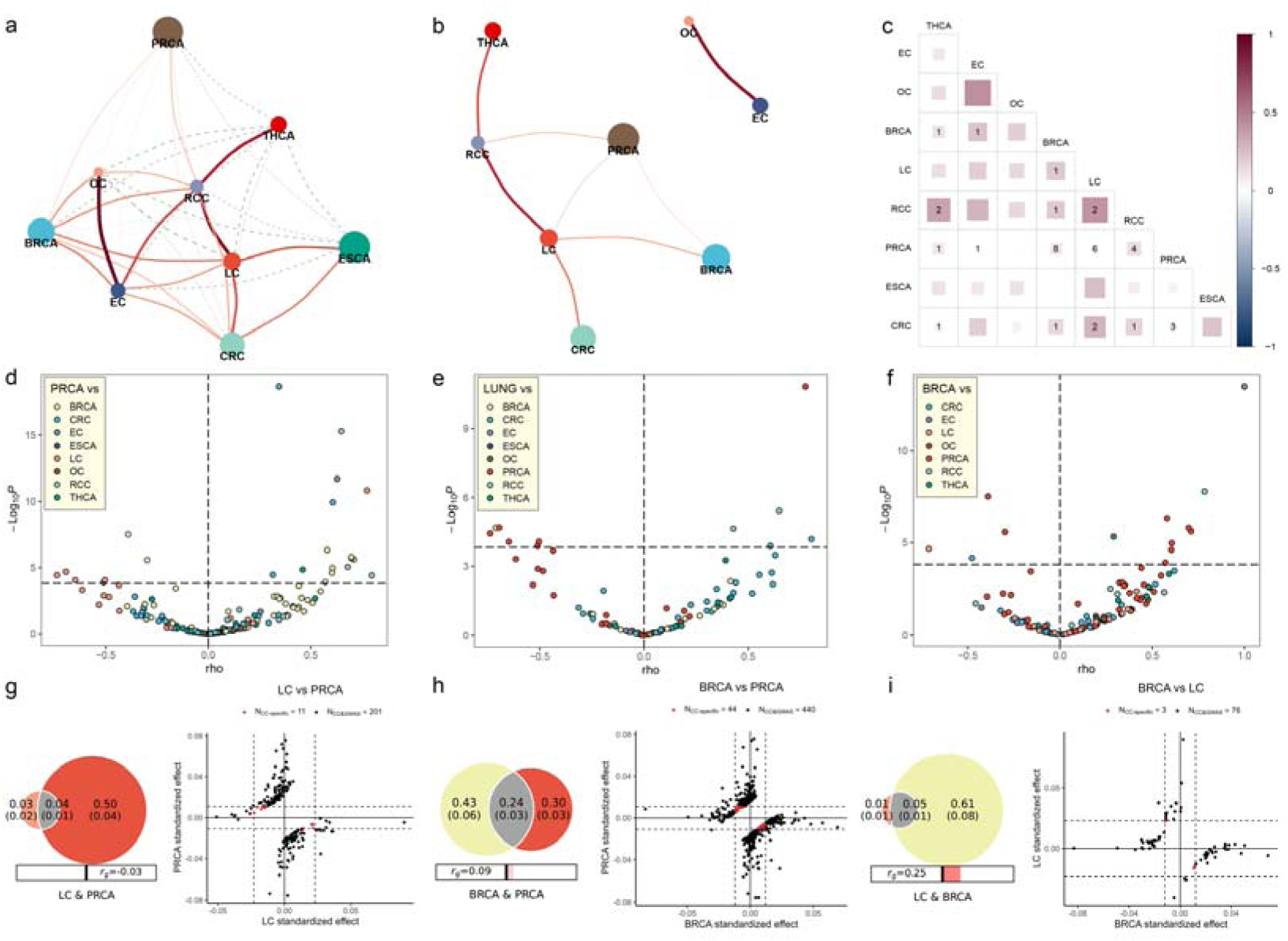
**Genetic correlations across nine carcinomas (genome-wide and local); pairwise polygenic overlap (MiXeR) and case–case contrasts by CC-GWAS.** (a) LDSC genetic-correlation network across nine carcinomas. Node size ∝ SNP heritability (ℎ^2^_SNP_) ; edge width encodes |r_g_|, color encodes direction and magnitude (red, positive; blue, negative). Edges not significant at FDRL<L0.05 are shown as gray dashedlines. (b) Conditional genetic network (GNA) showing partial genetic correlations after conditioning on the remaining traits. Visual encodings match panel a. (c) Lower-triangle heat map of pairwise LDSC r_g_ values; cell area and fill intensity scale with |r_g_| (red, positive; blue, negative; scale bar at right). Numbers in cells indicate the count of LD blocks with significant local genetic correlation from LAVA (Bonferroni-corrected across tested locus–pair combinations). (d–f) Local genetic correlations by LAVA for three cancers with the greatest extent of significant local sharing and pronounced antagonistic structure in pairwise scans: volcano plots anchored on (d) PRCA, (e) LC and (f) BRCA. Each point is an LD block; x-axis, local genetic correlation (p); y-axis, −log_10_P. Point color denotes the partner cancer (legend). Vertical dashed line marks p=0; horizontal dashed line marks the Bonferroni-corrected study-wide threshold across blocks. Positive p indicates concordant local effects; negative p indicates antagonistic effects. (g–i) Pairwise polygenic overlap (MiXeR) paired with CC-GWAS case–case contrasts for three representative cancer pairs: (g) LC vs PRCA, (h) BRCA vs PRCA and (i) BRCA vs LC. In MiXeR Venn diagrams, sectors encode the estimated numbers of trait-influencing variants unique to each cancer and shared (means with s.e., printed within sectors). Bars beneath Venns summarize genome-wide r_g_ . Right subpanels show standardized per-allele effects for independent loci from CC-GWAS contrasts; zero lines are shown; dashed lines indicate decision thresholds. Red points mark case–case–specific loci (significant in CC-GWAS but not in either case–control GWAS); black points mark loci detected in both CC-GWAS and case–control GWAS. Quadrants classify effect-direction concordance between cancers. Abbreviations as in Fig. 1.

### Revealing Large Polygenic Overlap and 145 Novel Independent Case–Case Loci Across Carcinoma Pairs

To quantify polygenic overlap that can be underestimated by genome-wide *r_g_* when effects are directionally mixed, we applied the bivariate causal mixture model (MiXeR) to five carcinomas with sufficient power as main analyses (Methods). Additional carcinomas with smaller Neff but acceptable univariate MiXeR fit were included as supplementary analyses. Across carcinoma pairs, we observed a wide spectrum of polygenic sharing. Even in low *r_g_* pairs, the overlap across carcinomas was extensive and exceeded what *r_g_* alone would suggest **(Figure S1; Table S3, S4)**. To localize variants driving cross-carcinoma differences, we then performed case–case GWAS (CC-GWAS)^14^. We identified 145 novel case–case loci with allele-frequency differences between carcinomas, each supported by directionally discordant effects across the paired cancers **(Fig. S2; Table S5)**, underscoring genetic mechanisms that differentiate related malignancies. Pairs involving PRCA, LC, and BRCA were most prominent because of the most negative regional genetic correlations and the high number of CC-specific loci identified **(Fig. 2d-i)**. Integrating local *r_g_* with CC-GWAS highlighted loci within the same genomic regions exhibiting directionally mixed effects across carcinomas, consistent with allelic heterogeneity across shared genomic regions.

### Multivariate Genetic Architecture of Carcinomas

We next used Genomic Structural Equation Modeling (Genomic SEM) to model shared genetic architecture across carcinomas, which is based on multivariable LD Score regression–derived genetic covariances and adjusts genetic overlap across distinct and even mutually exclusive samples^15,16^. In quality control, given uniformly low factor loadings in the exploratory factor analysis (EFA) and in leave-one-trait-out model estimates, PRCA was excluded, leaving eight carcinomas for analysis **(Table S6, S7)**. A combination of exploratory and confirmatory analyses prioritized two structures: a common factor model and a hierarchical model **(Fig. S3)**. The common factor model showed acceptable fit (χ² = 45.93, *P* = 8.20E-4; Comparative Fit Index (CFI) = 0.911; Standardized Root Mean Square Residual (SRMR) = 0.075; Akaike Information Criterion (AIC) = 77.93) **(Fig. 3a)**. All eight carcinomas loaded significantly on the common factor (P<5.0×10^−5), which accounted on average for 19.5% of genetic variance across carcinomas (range, 6.5% for ESCA to 40.6% for LC), with the remainder captured by trait-specific residual variance. A hierarchical model provided a superior fit (χ² = 26.36, *P* = 6.80E-2; CFI = 0.968; SRMR = 0.048; AIC = 64.36). Three correlated lower-order factors—F1 (respiratory/digestive; LC, ESCA, CRC), F2 (metabolic/endocrine; RCC, THCA) and F3 (hormone-related female reproductive; OC, EC, BRCA)—loaded onto a higher-order epithelial (e) factor **(Fig. 3b)**. Across traits, the hierarchical solution explained on average 33.8% of genetic variance (range: 9.3% for ESCA to 96.4% for RCC). Decomposition indicated a genome-wide general component (e-factor; mean 17.1%) plus domain-specific components (F1–F3; mean 16.7%). Illustratively, LC showed 59.5% explained variance (∼35.1% e and ∼24.4% F1), EC 34.3% (∼19.5% e and ∼14.8% F3), and RCC 96.4% (∼38.4% e and ∼58.0% F2), whereas ESCA and THCA retained predominantly trait-specific variance (9.3% and 13.0% explained, respectively). Together, these findings indicate that while a single dimension captures a modest portion of cross-carcinoma genetic signal, a hierarchical architecture—with a general epithelial factor and domain-specific factors—captures substantially more variance and clarifies the distribution of global versus domain-specific influences across carcinomas, paralleling patterns reported in other complex trait domains^17^.

**Fig. 3.**
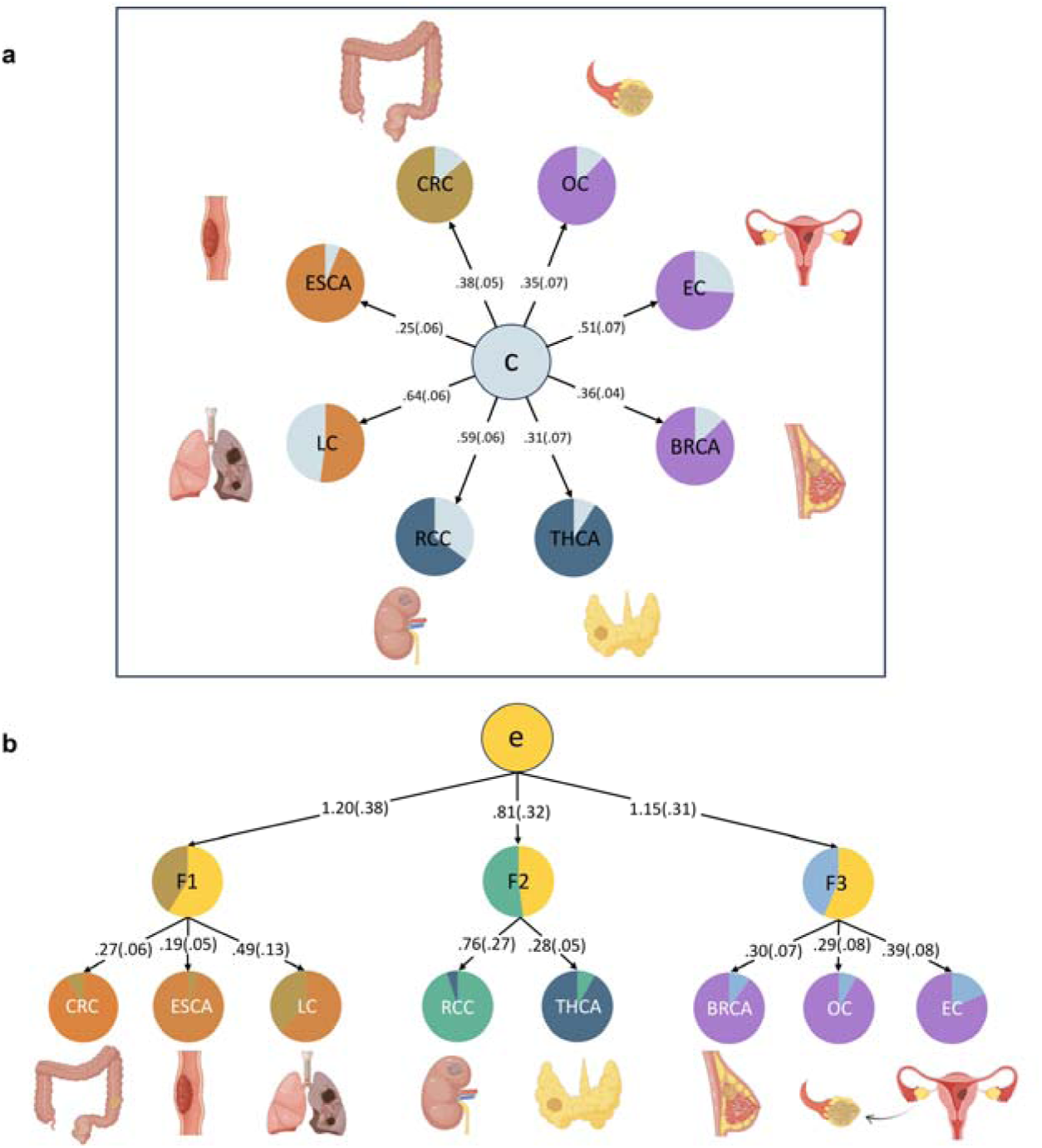
**Genomic SEM of shared genetic liability across eight carcinomas.** (a) Common factor model. A single latent factor (C) captures genetic covariation among LC, BRCA, ESCA, EC, OC, RCC, CRC and THCA. Numbers on arrows are unstandardized factor loadings with standard errors. Pie glyphs partition each trait’s SNP-heritability into variance explained by C (dark) versus trait-specific residual variance (light). The model was fit in Genomic SEM using diagonally weighted least squares (DWLS) on LDSC-derived genetic covariance and sampling-covariance matrices on the liability scale. (b) Hierarchical e-factor model. A higher-order factor (e) loads on three first-order factors mapping to carcinoma groupings: F1 → LC, ESCA, CRC; F2 → RCC, THCA; F3 →OC, EC, BRCA. Arrow labels are unstandardized path coefficients (s.e.) from e → F1–F3 and from each first-order factor to its cancers. Pie glyphs indicate variance explained by the parent factor (dark) versus residual variance (light). The e-factor model was fit under a fixed measurement model.

### Latent Factor Multivariate GWAS Identifies Pleiotropic Loci Related to Carcinogenesis

We subsequently used our hierarchical and common factor Genomic SEM models to perform latent factor multivariate GWAS, which leveraged GWAS summary statistics to estimate single-nucleotide polymorphisms (SNPs) associated with the latent factor **(Fig. 4)**. Variants showing heterogeneous cross-trait effects were excluded (P_QSNP_ < 5×10^−8). Loci were defined by LD clumping using a European reference panel (r^2<0.1; ±500Lkb; see Methods). Across the latent-factor GWAS, we identified 279 genome-wide significant pleiotropic loci (P<5×10⁻L). All 279 have been reported in single-cancer GWAS; however, 170 have not been reported as pleiotropic in prior cross-cancer scans^9–12^, indicating substantial pleiotropic novelty relative to earlier meta-analyses **(Tables S8–S9)**. The common factor and e-factor GWAS yielded 48 and 18 significant loci, respectively; both general latent-factor GWAS largely shared loci, were highly genetically correlated (*r_g_* = 0.894, SE = 0.057, P =2.39E-53) and each showed high *r_g_* (*r_g_*> 0.5) with the three-factor correlated first-order model GWAS **(Table S11)**, consistent with partly shared polygenic architecture that emphasizes distinct components of common liability. Each correlated factor exhibited moderate positive (>0.2) *r_g_* with its constituent cancers. As a quality-control diagnostic, we evaluated the attenuation ratio^18^ for each factor GWAS **(Table S10; Fig. S4)**. Estimates were near zero or negative for common factor, e-factor, F1, and F2 (approximately −0.04, −0.20, −0.02, and −0.27, respectively), and slightly positive for F3 (≈0.10 with confidence intervals spanning zero), indicating that residual confounding from population stratification or sample overlap is unlikely to materially affect the results. Negative ratios reflect intercepts <1—consistent with conservative test statistics—and may also arise from constraints inherent to the hierarchical specification (e.g., stronger regularization for factors defined by fewer contributing phenotypes), most notably for F2. To refine causal signals, we performed functionally informed fine-mapping. The 95% credible sets were obtained for 268 of 279 loci. Across these loci, 269 variants had posterior inclusion probability (PIP)>0.5, and 197 loci resolved to single-variant credible sets (PIP ≥0.95) **(Table S12)**. These results nominate a compact set of putative causal variants for downstream functional interrogation. To investigate the cell-type-specific enrichment, we mapped latent-factor GWAS signals onto a pan-carcinoma single-cell RNA-seq atlas comprising 33 cancer types. Each latent-factor GWAS showed significant enrichment in malignant epithelial cells, although lower-order factor GWAS revealed pronounced heterogeneity within these enriched epithelial populations **(Fig. 4; Fig. S5)**. Together, these findings indicate that the latent-factor framework captures both shared and group-specific genetic effects across carcinomas.

**Fig. 4.**
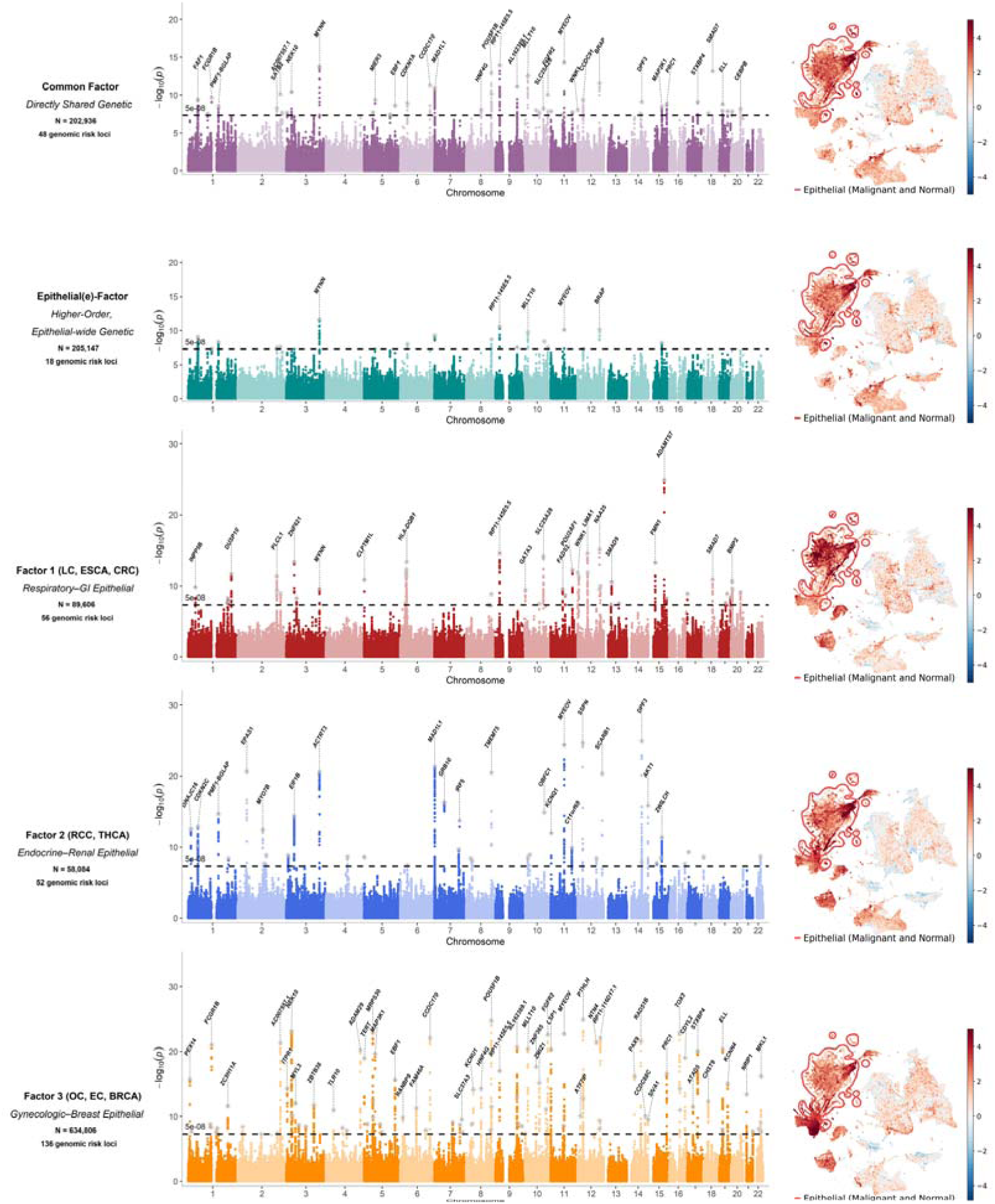
Manhattan plots and cell-type enrichment of latent factor multivariate GWAS. Panel shows –log10(P) for SNP associations across autosomes (Chromosomes 1–22); the dashed line marks genome-wide significance (P = 5 × 10⁻L). Lead variants at genome-wide significant loci are annotated by nearest gene. Panel titles report the effective sample size (N) for each factor GWAS and the number of independent genomic risk loci. UMAP plots are colored by scDRS scores computed from the latent-factor GWAS and pan-carcinoma single-cell landscape, with red indicating higher disease relevance. Epithelial cells, including both malignant and normal subtypes, are highlighted with a red outline and shown in the inset.

### Genetic Overlap Between Latent Factors and Carcinoma Traits, and Cross-Ancestry Transferability

To assess whether the latent factors track distinct carcinoma biology, we estimated genetic correlations between each latent-factor GWAS and 15 TCGA-annotated carcinomas spanning diverse tissues **(Fig. 5; Table S13)**. For both the common factor and the e-factor, most carcinomas exhibited moderate-to-high *r_g_*, consistent with broad yet graded sharing across epithelial cancers. In contrast, prostate cancer showed uniformly low *r_g_* with all latent factors, mirroring its discordant local effects and indicating a distinct genetic architecture relative to other carcinomas. We next evaluated cross-ancestry generalizability. Using large East-Asian (EAS) population GWAS for BRCA, PRCA, CRC and gastric cancer (STAD), we constructed an EAS common factor GWAS under the same QC and modeling pipeline and tested trans-ethnic genetic correlations with the European (EUR) latent factors **(Table S14; Fig. S6)**. The EAS common factor model demonstrated excellent fit (χ²=0.889, P=0.641, AIC=16.9, CFI=1.00, SRMR=0.025) and showed substantial cross-ancestry correlation with the EUR common factor (ρg=0.71, SE=0.08, P=2.85E-04) and with the EUR e-factor GWAS (ρg=0.78, SE=0.10, P=2.17E-02) **(Fig. 5)**. These results support the preliminary evidence of consistency across populations of the pan-carcinoma latent-factor architecture.

**Fig. 5.**
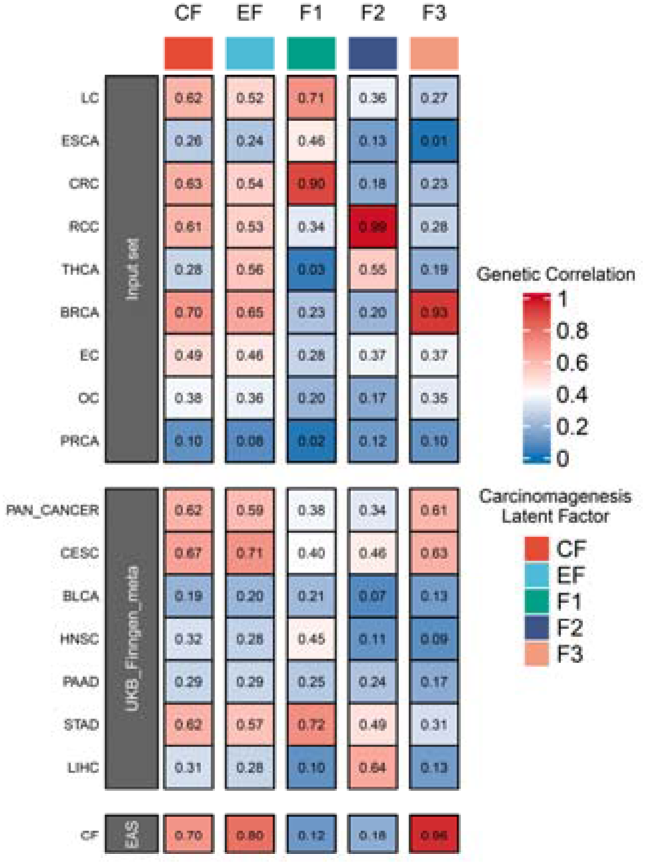
**Genetic correlations between latent factor and carcinoma GWAS.** Heatmap of bivariate genetic correlations (LDSC rg between five factor GWAS—common factor (CF), higher-order epithelial factor (EF) and first-order factors F1–F3—and external carcinoma GWAS panels (input cancers, additional UKB/FinnGen carcinomas including a pan-cancer meta-analysis, and an East-Asian common factor). Cell color and size reflect |rg| (scale at right); values are point estimates. Higher values indicate stronger shared genetic liability. Additional abbreviations for this figure: PAN_CANCER, pan-cancer meta-analysis; CESC, cervical cancer; BLCA, bladder cancer; HNSC, head and neck cancer; PAAD, pancreatic cancer; STAD, stomach cancer; LIHC, liver cancer; EAS CF, East Asian common factor. Other abbreviations as in Fig. 1.

### Multi-trait Multivariate GWAS Identifies Novel Loci Related to Carcinogenesis

To increase discovery power for novel loci, we applied a multi-trait multivariate GWAS^19^ within each latent-factor cluster, thereby boosting the trait-specific effective sample sizes of the constituent single-cancer GWAS. For the MTAG results, per-trait maxFDR values ranged from 0.001 to 0.19; for most traits the cluster-based MTAG reduced the baseline maxFDR **(Table S15)**. Compared with the original single-trait GWAS, MTAG increased the estimated effective sample sizes and raised mean χ² values; LDSC analyses indicated that, for cancers with smaller original sample sizes, estimated heritability generally increased after the cluster-based multi-trait multivariate GWAS, consistent with improved power to detect genetic signal; we report h² estimates and LDSC intercepts to exclude inflation from confounding or sample overlap **(Table S15)**. Using these MTAG results we identified 44 novel loci across five cancer types **(Fig. S7; Table S16)**. Given the intermediate maxFDR for some traits, we sought replication in an independent smaller cohort and performed pairwise cross-cancer replication analyses; five of the MTAG-identified novel loci replicated **(Table S16–S17)**. The lead SNPs included rs4513875 (7p22.3, MAD1L1, P = 1.73E-10) for THCA; rs147144681 (15q25.1, CHRNA5, P = 2.51E-15) for ESCA; and rs1696840 (10q26.13, FGFR2, P = 6.21E-13), rs3803661 (16q12.1, TOX3, P = 1.29E-09) and rs657686 (11q13.3, CCND1, P = 1.63E-08) for ENDO. These results suggest that biologically plausible cluster-based multivariate GWAS can improve detection of genetic signals.

### Integrated Multi-omics Evidence Prioritizes Pleiotropic Effector Genes Underlying Carcinogenesis

We next sought to nominate pleiotropic effector genes that mediate cross-carcinoma risk. We integrated (i) functionally informed fine-mapping and annotation-based gene prioritization, (ii) cross-tissue transcriptome-wide associations based on SEM framework, and (iii) plasma proteome-wide associations based on SEM framework. This multi-layer framework yielded 337 candidate pleiotropic genes **(Tables S18–S20, S25)**. To further prioritize effectors with convergent biological support, we overlaid multi-omics readouts restricted to signals recurrent across carcinoma types within each modality: bulk RNA-seq differentially expressed genes significant in ≥2 carcinoma entities (|logFC|>1, FDR<0.01), pan-cancer proteomics proteins enriched in ≥2 carcinoma types (logFC>1, FDR<0.01), and robust carcinoma program markers from scRNA-seq replicated across >2 tissues, alongside tumor microenvironment (TME) markers (top 10 genes) spanning 9 major cell classes and 146 subtypes **(Tables S21–S23)**. Notably, most candidate genes are enriched in malignant rather than TME cells, indicating that their carcinogenic effects are mediated primarily through cell-autonomous pathways rather than microenvironmental pathways **(Fig. 6)**. Genes supported by ≥3 independent evidence streams and at least one genetic evidence were designated high-confidence pleiotropic effectors, yielding 167 prioritized genes **(Fig. 6, Tables S25)**. Pathway enrichment of these genes highlighted epithelial and neoplastic programs **(Fig. S8)**, with top terms including WikiPathways: Cancer pathways (Padj =1.18E−8), WikiPathways: Head and neck squamous cell carcinoma (Padj =1.18E−8), GO Biological Process: epithelial cell proliferation (Padj=2.75E−7), GO Biological Process: epithelial cell differentiation (Padj =5.37E−7), and in utero embryonic development (Padj =3.98E−6). Collectively, these analyses converge on a compact set of effector genes enriched in epithelial growth and differentiation pathways that plausibly underpin shared carcinogenic mechanisms.

**Fig. 6.**
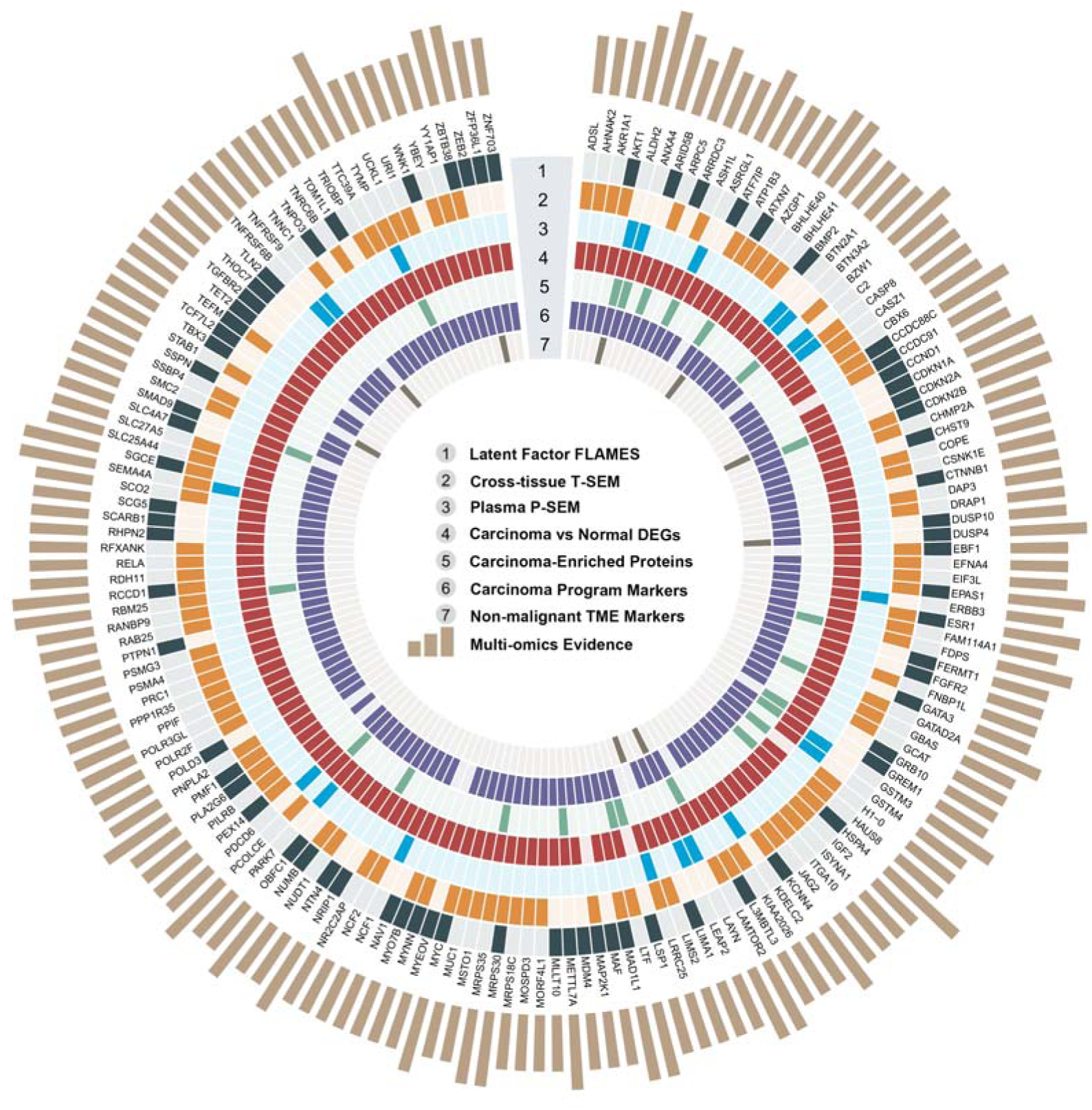
**Multi-omics prioritization of pleiotropic effector genes for Genomic SEM factors.** Radial heatmap summarizing convergent evidence for 167 high-confidence pleiotropic effector genes implicated by the factor GWAS. Spokes correspond to genes (labels around the circumference). Colored tiles indicate significant support from a given evidence layer; empty tiles indicate no support. From center outward, rings encode: (1) *Latent-factor FLAMES* (fine-mapping/annotation–based gene prioritization at factor loci); (2) *Cross-tissue T-SEM* (transcriptome-wide associations aggregated across tissues modeled with Genomic SEM); **(3)** *Plasma P-SEM* (protein-level associations modeled with Genomic SEM); (4) *Carcinoma vs normal DEGs* (bulk RNA-seq differential expression genes); (5) *Carcinoma-enriched proteins* (proteomic enrichment in tumor); (6) *Carcinoma program markers* (membership in curated single-cell epithelial-carcinoma programs); (7) *Non-malignant TME markers* (markers across 9 main cell types and 146 related subtypes in tumor-microenvironment). Outermost beige bars give an integrated multi-omics evidence score per gene (higher bars = more supporting layers), used to rank candidates; top-ranked genes are annotated at the perimeter.

### Phenome-wide Causal Associations with Carcinogenesis

To delineate putative causal risk pathways^20^, we performed phenome-wide Mendelian randomization (PWMR) using the latent-factor GWAS as outcomes and 1,649 non-cancer phenotypes from an independent European-ancestry cohort (N=451,206) as exposures. Instruments were clumped from genome-wide significant variants; effects were estimated primarily by inverse-variance weighting (or Wald ratio for single-IV exposures), with concordance required across sensitivity estimators **(Fig. S9)**. Study-wide significance was set at α=6.06×10^−6. For the general latent factor, PWMR recapitulated broad epidemiologic signals—including higher BMI, body weight, height, circulating pro-BNP, chronic airway obstruction, and actinic keratosis—implicating systemic metabolic and epithelial stress pathways in cross-carcinoma risk **(Fig. 7; Tables S26)**. Factor-specific scans identified traits aligned with the domain structure: for F1 (respiratory/digestive), colon polyps (OR=1.59, SE=0.16, P=2.29E-6) and chronic airway obstruction (OR=1.26, SE=0.04, P=8.38E-13); for F2 (metabolic/endocrine), obesity (OR=1.22, SE=0.34, P=1.01E-11); and for F3 (hormone-related female reproductive), maternal history of breast cancer (OR=1.65, SE=0.06, P= 6.20E-50) and other non-malignant breast conditions (OR=1.14, SE=0.01, P=1.89E-34). These phenome-wide findings both reproduce known risk architectures at the level of individual cancers and generalize them to latent dimensions that capture shared and domain-specific carcinogenic liability.

**Fig. 7.**
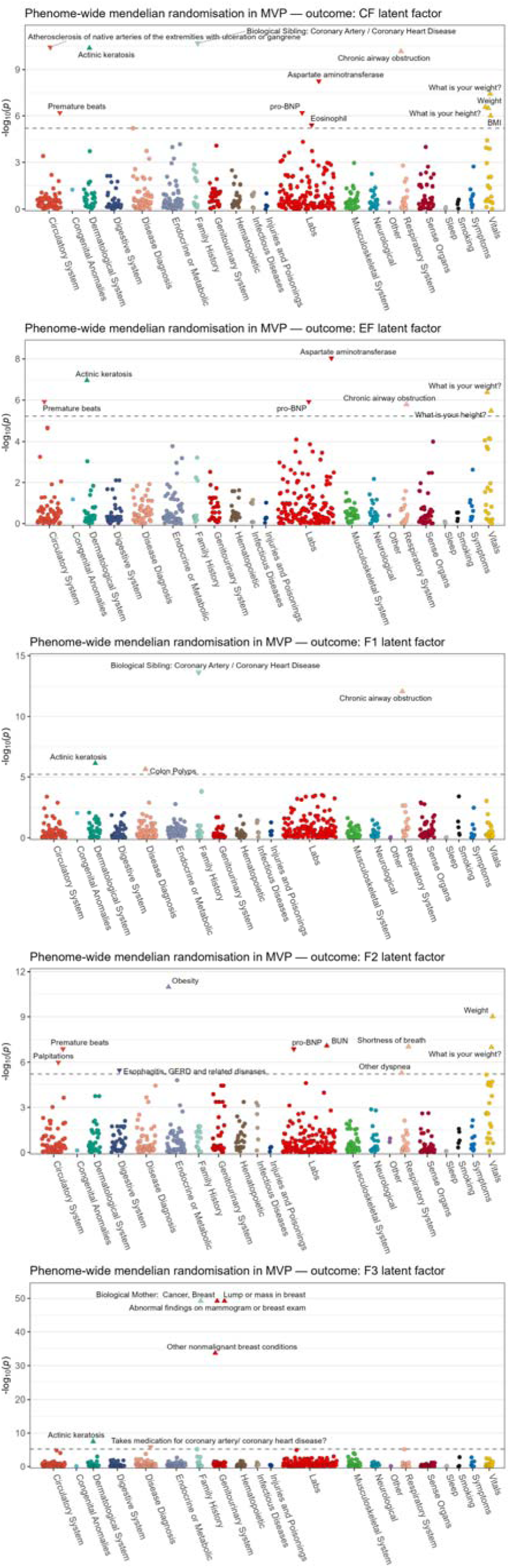
**Phenome-wide Mendelian randomization (PWMR) of MVP traits on Genomic SEM factors.** Panels (top to bottom) show outcomes common factor (CF), e-factor (EF), F1 (LC–ESCA–CRC), F2 (RCC–THCA) and F3 (OC–EC–BRCA). The x-axis groups 1,649 MVP phenotypes by 21 categories (points jittered within groups); the y-axis is −log₁₀(P). The dashed line marks the Bonferroni-corrected 5% threshold across traits. Point color indicates MVP category. Marker shape encodes significance and effect direction: circles, non-significant; upward triangles, significant positive; downward triangles, significant negative. Selected traits exceeding the threshold are labeled.

## Discussion

Our analyses map the shared and differentiated genetic architecture of nine etiologically and clinically distinct carcinomas. Several main findings are consistent across complementary methods. First, we confirm pervasive cross-carcinoma genetic overlap. By triangulating evidence at the genome-wide, regional, and locus levels, we uncover widespread mixtures of concordant and discordant effects between carcinoma pairs, implying that prior work likely underestimated cross-cancer genetic correlation by averaging over directionally opposed signals. Second, using genomic structural equation modeling, we capture across carcinomas shared pleiotropy with two general factors, both a single common factor and a hierarchical higher-order model. The common factor reflects a broad, transdiagnostic liability, whereas the hierarchical solution partitions eight carcinomas into three biologically coherent groups and summarizes their shared signal with a higher-order factor (the e-factor). On average, these latent structures explain a substantial proportion of SNP heritability across individual carcinomas (19.5% for the common factor; 33.8% for the hierarchical factor). Both models recapitulate moderate-to-high genetic correlations across most carcinomas and show transferability across ancestry groups. Third, leveraging GWAS of the latent factors, we identified 279 pleiotropic loci across 103 regions, including 170 loci not previously reported in cross-cancer scans. In addition, multivariate analyses increased discovery power in single-cancer GWAS, yielding 44 novel loci—five of which were successfully replicated. Fine-mapping and functional genomic integration nominated 337 likely effector genes; multi-omics prioritization then produced 167 high-confidence pleiotropic effector genes that are enriched for epithelial growth and differentiation programs, nominating targets with potential utility for broad anticancer strategies while helping anticipate mechanism-based adverse events.

Integration across analytic granularities clarifies how cross-cancer overlap arises. The modest genome-wide correlations are in line with earlier cross-cancer observations^9–11^, but our conditional and local analyses show that such averages can understate true sharing when effects differ in direction. Loci with opposing allelic effects across cancers frequently colocalize with regions of significant local correlation, indicating that allelic heterogeneity within shared neighborhoods is common^21^. The 5p15.33 region harboring *TERT* exemplifies how pleiotropy and cross-cancer differentiation can coincide^22^. Such architecture may help explain why targets extrapolated across indications sometimes fail in pan-cancer settings ^23,24^. Together, these results argue that interpreting cross-cancer genetic architecture requires moving beyond global correlation to conditional and locus-resolved perspectives that can accommodate antagonistic structure.

The multivariate latent structure provides a unified, principled framework anchored in summary-statistics-based genetic covariance^15,16^. A common factor captures broad epithelial liability, but a hierarchical latent factor solution offers superior fit and biological interpretability: F1, comprising respiratory and digestive tract carcinomas, aligns with evidence for extensive genetic sharing across these systems in diverse populations^25^. F2 groups thyroid and renal cancers, consistent with bidirectional associations arising from shared genetic and environmental risks^26^. F3 comprises three carcinomas of female reproductive organs, whose epithelial compartments are hormone-responsive and enriched for estrogen-related oncogenic processes; this grouping echoes a gynecologic iCluster observed in prior pan-cancer multi-omics analyses^4^. The e-factor GWAS exhibits broad genome-wide sharing with each domain-specific factor as well as with the common factor, pointing to a shared epithelial component that is complemented by additional, domain-specific liabilities.

Applying multi-trait multivariate GWAS within this latent structure substantially increases effective sample sizes for constituent single-cancer GWAS, improves the proportion of heritability explained, and facilitates identification of novel loci that are difficult to detect in univariate scans. For example, we detected an association between ESCA risk and a locus at 15q25.1 (lead variant rs55853698) within the CHRNA5 gene. This association is biologically plausible, given that smoking is a well-established risk factor for ESCA. The CHRNA5 gene, which encodes the α5 nicotinic acetylcholine receptor subunit and influences brain reward circuitry^27^, has been previously implicated in both smoking behavior and lung cancer^28,29^. Separately, we identified a germline association for OVCA at FGFR2 (lead variant rs2981582). Notably, FGFR2 is a recurrent somatic driver in endometrial carcinoma, where activating alterations occur in a substantial subset of cases^30^.

There are several limitations. First, although the carcinomas analyzed cover a large share of incident cancers and yielded a general factor with moderate-to-high genetic correlations across sites, many carcinoma types were not included. Given the complex cross-tumor clustering observed in RNA-seq data, future work should incorporate additional large-scale carcinoma GWAS and adopt more flexible hierarchical models to better capture pleiotropy. Second, the factor models were constructed from LDSC-derived genetic covariance, which can attenuate signal at loci that combine concordant and discordant effects. We excluded PRCA, which showed pronounced negative genetic correlations with other traits during model construction, and removed SNPs showing significant heterogeneity in the Q-test; this choice may limit the completeness of the inferred factor structure. Consistent with the CC-GWAS results, pairwise differences between carcinomas appear to be driven largely by directional heterogeneity at specific loci, indicating that locus-based models will be needed for more fine-grained insight. Third, due to the limited availability of GWAS in non-European ancestry groups, analyses focused primarily on EUR populations. We did construct a common factor in an EAS sample by combining the four largest carcinoma GWAS and observed high cross-ancestry genetic correlation with the EUR model; however, the small set of carcinoma types and relatively modest sample sizes reduce power to fully characterize the complex genetic architecture in EAS populations. Finally, although we performed an independent replication analysis to confirm loci identified in the multi-trait analysis, the largest discrepancy in effective sample size reached fourfold between the replication and original GWAS cohorts; this imbalance may have substantially underestimated the number of novel genomic loci.

In conclusion, we systematically dissect the shared and cancer-specific genetic architecture of carcinomas at varying levels of genetic granularity by triangulating evidence from cutting-edge statistical genetic and functional genomic analyses, which reveal previously underestimated genetic overlap. We introduce the first hierarchical genomic latent model of carcinomas and demonstrate its added potential to explain shared genetic structure across cancers and illustrate its utility for identifying novel pleiotropic loci. Our study may be helpful for developing novel preventive and therapeutic strategies that minimize the broad range of side effects in cancer therapy.

## Methods

### Data preparation

We assembled the largest available European-ancestry GWAS summary statistics for nine carcinoma phenotypes, spanning a total of 429,158 cancer cases. The cancers included breast cancer (BRCA)^31^, colorectal cancer (CRC)^32^, endometrial cancer (EC)^33^, esophageal cancer (ESCA)^34^, lung cancer (LC)^35^, ovarian cancer (OC)^36^, thyroid cancer (THCA)^37^, prostate cancer (PRCA)^38^, and kidney cancer (RCC)^39^. We harmonized and filtered each GWAS dataset following established protocols from previous studies^9–11^: each GWAS summary required a heritability |Z| value greater than 4 to ensure high credibility for downstream genetic analyses^40^. Variant genomic coordinates were lifted over to GRCh37 and rsIDs were updated using dbSNP build 156; variants with minor allele frequency (MAF) ≤ 1%, imputation INFO score ≤ 0.9, extreme effect size (|β| ≥ 3), or very small standard error (SE ≤ 1.00E-5) were removed. We calculated Neff for each study using the formula recommended by GenomicSEM^15^, which more appropriately accounts for case–control ascertainment differences in large meta-analytic GWAS^41^. These Neff values were used in downstream analyses to ensure consistent weighting across cohorts.

### Genetic correlation network analysis

We estimated genetic correlations between each pair of cancers using multivariate LD Score regression as implemented in GenomicSEM v0.05^15,16^. This analysis was restricted to HapMap3 SNPs and used LD scores computed for the 1000 Genomes (1KG) Project Phase 3 European population, excluding the extended major histocompatibility complex (MHC) region. We excluded any trait with a SNP-heritability Z-score ≤ 4, as such low-heritability traits can yield unstable genetic correlation estimates^40^. We converted SNP-heritability to the liability scale assuming a sample prevalence of 0.5 and a population lifetime prevalence for each cancer based on SEER non-Hispanic white population data (seer.cancer.gov). Using the genetic covariance matrix from LDSC^16^, we then applied the traitNET function of the Genomic Network Analysis (GNA) v0.01^42^ to construct a network of conditional genetic associations among the cancers. This approach estimates a sparse precision matrix from the genetic covariance, yielding a trait network in which edges represent direct (conditionally independent) genetic correlations between cancers, after accounting for indirect sharing through the broader network. Significance for network connections and other genome-wide analyses was assessed at a FDR < 0.05.

### Local genetic correlation analysis

To further dissect genetic sharing across specific genomic regions, we employed complementary local genetic correlation methods. First, we applied the bivariate causal mixture model approach in MiXeR^43^ to quantify polygenic overlap between traits. We followed the MiXeR v1.3 pipeline by using the provided sumstats.py script to format and harmonize summary statistics (excluding the MHC region to avoid bias from complex LD) and including only traits AIC>0 and Neff>50,000 in univariate MiXeR to ensure adequate power^44^. We fit univariate mixture models for each trait and subsequently bivariate mixture models for each included cancer pair to estimate the number of shared causal variants. Each analysis was run 20 times, and the results were averaged to obtain more robust estimates. All model fits met recommended criteria (e.g., best_vs_min_AIC>0 and best_vs_max_AIC>0), indicating that the mixture models provided a good description of the data. Linkage disequilibrium information for MiXeR was derived from the combined UK10K–1KG Phase 3 European reference panel (N = 4,285)^45,46^.

We next performed local genetic correlation analysis using LAVA v0.1.0^47^. For LAVA, we entered the GWAS summary statistics (aligned to a common set of SNPs and alleles) along with an estimate of any sample overlap between traits (as estimated by LDSC intercept attenuation). We used an external European-ancestry LD reference panel from UK Biobank (N = 100,000) and predefined 2,495 approximately independent genomic loci (covering the whole genome) for partitioned analysis. As a quality control step, we first conducted univariate LAVA heritability tests for each locus and each trait, and we excluded loci with non-significant heritability (P > 2.00E-5, corresponding to 0.05/2,495). We then tested for local *r_g_* between every pair of cancers within each remaining locus. In total, 332 locus-by-pair *r_g_*tests were performed, so we applied a Bonferroni-corrected significance threshold of P < 1.51E-4 (0.05/332) for declaring significant local rg results.

### Pairwise Case–Case Differential GWAS

To uncover the signals that differ in direction between carcinomas, we conducted CC-GWAS, which is a case–case association test of two different traits to detect loci with allele-frequency differences based on their respective case-control GWAS data^14^. In this framework, a weighted contrast of case–control GWAS summary statistics is estimated, so that cross-carcinoma individual-level matching is not required. Two complementary tests are integrated. CC-GWAS_OLS, derived from analytic expectations of case–control genetic differences for each disease, is tuned to maximize power while controlling type I error at variants null for both disorders. CC-GWAS_Exact is used to control type I error at “stress-test” variants that affect both disorders but yield no case–case frequency difference. A variant is declared associated with case–case status only when the OLS test attains genome-wide significance and the exact test yields P < 1×10^−4; the number of stress-test variants is upper-bounded because these represent causal sites. To limit false positives caused by subtle differential tagging of stress-test variants—even within a single ancestry—specific filters are applied. The input m was set to the average number of independently associated SNPs between the two traits^48^.、

### Genomic structural equation modeling

Structural equation modeling (SEM) was used to identify latent dimensions of genetic covariance among the cancers. Genomic SEM extends traditional SEM by using the genetic covariance matrix (S) derived from GWAS summary data to model common factors underlying multiple traits. We constructed a genomic SEM of the nine cancers, based on the LDSC genetic covariance matrix S (and its sampling covariance matrix V) estimated as described above. The sum of effective sample sizes for each trait was included in the covariance estimation^41^. Prior to factor analysis, we adjusted the covariance matrix to be positive-definite using a nearest positive definite algorithm (nearPD), yielding a smoothed matrix Ssmooth. We empirically determined the optimal number of latent factors using multiple criteria. First, we performed parallel analysis on Ssmooth with 100 random permutations using the paLDSC() function in GenomicSEM^49^, comparing the observed eigenvalue distribution to that expected under a null model. We also applied three scree test methods on the genetic correlation matrix: the Kaiser criterion^50^, the acceleration factor method, and the optimal coordinates method^51^. For the set of nine cancers, the parallel analysis, acceleration factor, and optimal coordinates all suggested a single common factor, whereas the Kaiser criterion suggested up to three factors. We next conducted EFA to further probe the factor structure. We used the R factanal function with promax rotation (allowing factors to correlate), testing both a one-factor model and a three-factor model. Because PRCA showed very low factor loadings (<0.2) in these analyses, we conducted leave-one-trait-out Genomic SEM modeling to investigate which traits should be excluded; in each analysis, one carcinoma trait was excluded and the remaining eight were used to estimate the common factor model. Based on model-fit comparisons, we removed the PRCA and repeated the EFA with (number of factors = 1 or 3) on the remaining eight cancers. In the three-factor EFA of eight cancers, each factor had at least one cancer with a high loading (> 0.5). Given that the three factors were still substantially intercorrelated, we specified a hierarchical (second-order) model comprising three first-order factors and a higher-order factor interpreted as a general epithelial carcinoma factor (“e-factor”) capturing variance shared across epithelial cancers. In the hierarchical specification, covariances among first-order factors were fixed to zero (orthogonal conditional on the higher-order factor); where biologically prespecified, we allowed targeted residual correlations between specific first-order factors. Models were fit in GenomicSEM (DWLS) using LD score–derived genetic covariance matrices, and model fit was evaluated using standard indices, considering CFI > 0.90 and SRMR < 0.08 as indicating acceptable fit.

### Latent factor multivariate GWAS

To identify genomic loci associated with the latent factors, we carried out common factor GWAS and multivariate GWAS using GenomicSEM. We first used the GenomicSEM sumstats function to align alleles across studies and to standardize SNP effect sizes and SEs (rescaling each to unit variance of the phenotype). After removing SNPs that led to non-positive-definite covariance matrices (likely due to problematic variant annotations), we conducted our analysis on a final set of approximately 2.50 million SNPs present in all eight cancer GWAS. We then used the commonfactorGWAS function to estimate SNP associations with the common factor, and the userGWAS function to estimate SNP associations with the e-factor and each specific factor in the three correlated factors. We filtered out any SNP that produced improper solutions (e.g., negative variance estimates or non-convergent covariance matrices) or that showed significant heterogeneity in their effects across traits (QSNP p-value < 5×10^−8), as such variants violate the assumption of a single shared effect. As a quality control, we calculated the LDSC attenuation ratio^18^ for each latent factor GWAS. Finally, we estimated the effective sample size of each factor GWAS using the method of Mallard et al.^52^, restricting to SNPs with MAF 10–40% to obtain stable estimates.

### Multi-trait multivariate GWAS and replication

Multi-trait analysis of GWAS (MTAG) is a generalized, inverse-variance-weighted meta-analysis method that utilizes genetic covariance between traits to boost statistical power^19^. Considering the genetic correlations and biological relationships, traits within each latent factor cluster were input separately to minimize the influence of directionally discordant effects. The cross-trait and single-trait upper bound of the false discovery rate (maxFDR) was calculated to evaluate the extent of overall inflation. We conducted a replication analysis to verify the identified genomic loci using independent cohorts, including corresponding cancer GWAS summary statistics from the Million Veteran Program (MVP) ^53^ and a meta-analysis of FinnGen and the UK Biobank^54,55^.

### Defining Risk Loci and Fine-Mapping of Causal Signals

We first identified independent genome-wide significant loci in both CC-GWAS and multivariate GWAS using linkage disequilibrium clumping in PLINK v2.0^56^ (index SNP P < 5×10^−8, clumping r^2 < 0.1, window size ± 500 kb). Overlapping loci were merged into a single region. We then compared the identified loci with the participating cancer GWAS and pleiotropic loci defined by previous cross-cancer meta-GWAS studies^9,11^. The loci were considered novel when the lead SNP was more than ±1 Mb away and r^2 < 0.1 with any previously identified genome-wide significant pleiotropic SNP (P < 5×10^−8). To pinpoint likely causal variants from the factor GWAS results, we carried out multi-step fine-mapping analyses. For each locus, we applied the Bayesian fine-mapping method CARMA (Causal Robust Mapping with Annotations)^57^ to compute 95% credible sets of causal variants, incorporating 263 functional annotations from baselineLD v2.2^58^ as priors to improve causal variant identification, each locus was expanded by ±100 kb to capture the remaining LD structure. We report CARMA 95% credible sets (ρ=0.95) allowing up to K=10 causal variants per locus; multiple variants within a credible set can exhibit high PIPs when signals are correlated or multi-causal, as expected under CARMA’s construction. For loci without enough SNPs for CARMA, we additionally performed approximate Bayes factor (ABF) fine-mapping, which is a conservative fine-mapping method based on Bayes factors and assumes up to one causal variant per locus^59^. We utilized the combined UK10K–1KG EUR reference panel for LD calculations^45,46^. Prior to fine-mapping, we used the susie_rss function from the SuSiE package^60^ to align alleles and remove any SNPs with mismatched alleles between the summary statistics and reference panel, ensuring a consistent LD matrix.

### Cell type-specific enrichment analysis

To assess cell-type enrichment of genetic signals from the latent-factor GWAS, we applied single-cell disease relevance score (scDRS, v1.0.4) analyses^61^. Precomputed pan-cancer meta-cells derived from a single-cell atlas covering 33 carcinoma types and 16 annotated cell types were used^62^. Gene-level P-values and Z-scores were obtained from the latent-factor GWAS summary statistics using MAGMA v1.10^63^. For each cell in the scRNA-seq atlas, scDRS aggregated expression of GWAS-implicated genes to compute a disease score and generated 1,000 Monte-Carlo control scores from matched random gene sets; scores were then normalized and converted to cell-level P-values. We ran scDRS’s compute_score with default parameters. Finally, we tested cell-type-level associations to identify broad cell types associated with disease status and to quantify within-type heterogeneity across individual cells. Reported multiple testing corrected P-values.

### Cross-cohort and cross-ancestry validation

We evaluated the generalizability of the latent factors across different cohorts and ancestries. First, we calculated genetic correlations between each latent factor GWAS and each of the original nine cancer GWAS using LDSC^16^. A moderate-to-high genetic correlation (rg > 0.2) was interpreted as evidence that the latent factor captured a substantial portion of the genetic signal of that cancer trait. Next, we tested for genetic correlation between the latent factors and different cancer GWAS from external cohorts. We considered eight cancer GWAS that can be annotated with TCGA cancer code from a recent meta-analysis of FinnGen and UK Biobank^54,55^, as well as a pan-cancer meta-analysis spanning 909,373 individuals (187,278 cases and 722,095 controls). Finally, we constructed a latent common factor using four large East-Asian-ancestry GWAS (for breast, stomach, colorectal, and prostate cancer), applying the same QC and analysis pipeline as in the European data, and we assessed its similarity to the European factor via cross-ancestry genetic correlation using Popcorn^64^ with precomputed 1KG EUR and EAS LD scores.

### Transcriptome-wide and Proteome-wide SEM

We employed transcriptome-wide structural equation modeling (T-SEM)^65^ to identify putative gene targets underlying the common cancer factor. T-SEM integrates multivariate LDSC results with gene-based association signals from transcriptome-wide association studies (TWAS) to test each gene’s effect on a latent factor. We first performed gene-level TWAS for each cancer trait using FUSION^66^, imputing gene expression based on cis-expression quantitative trait loci (cis-eQTL) weights. We used 3 cross-tissue weights generated in GTEx v.8 by sparse canonical correlation analysis (sCCA) across 49 tissues^67^ and the merged UK10K–1KG LD reference panel^45,46^ for these TWAS analyses. We then combined the TWAS results from all traits in a T-SEM analysis to identify genes whose predicted expression showed a significant association with the latent factor.

Analogously, we developed a proteome-wide SEM (P-SEM) approach to find shared protein influences on the cancer factor, which may be helpful for developing cross-carcinoma biomarkers. This approach mirrors T-SEM but uses protein-level association data from proteome-wide association studies (PWAS), thereby highlighting potential druggable targets^68^. We performed PWAS using the recently introduced BLISS method^69^, which enabled summary-statistics-based testing of protein–trait associations. We leveraged three available European plasma cis-pQTL weight sets (total N ≈ 92,446) that measure up to 5,769 proteins and corresponding LD references for these analyses. The models for 2,129 proteins with estimated heritability exceeding 0.01 were included. The resulting PWAS signals across carcinomas in all latent factor models were then analyzed in the Genomic SEM framework (P-SEM) to identify proteins significantly associated with the latent factor. We applied a Bonferroni-corrected significance threshold of P < 4.70E-6 (0.05/2129/5) for P-SEM and P < 2.64E-7 (0.05/37920/5) for T-SEM.

### Functional Annotation of Genetic Variants

To prioritize likely effector genes at associated loci, we applied the Fine-mapped Locus Assessment of Multiple Effector genes (FLAMES) v 1.1.2 tool^69^. FLAMES evaluates each gene’s candidacy by integrating fine-mapped variant-to-gene links with additional evidence of gene function or convergence across datasets. We supplied FLAMES with the 95% credible set variants from our fine-mapping results and with independent gene prioritization results from Polygenic Priority Score (PoPS) v0.2^70^. Genes above the cumulative 75% precision threshold were considered high-confidence effector genes driving the association signals^71^.

### Multi-omics Gene Prioritization and Enrichment

To highlight robust pan-carcinoma biological targets, we intersected our multi-level results with external molecular data. We included multi-omics evidence as follows: 1. Differentially expressed genes (DEGs) based on bulk RNA-seq from GEPIA2, 22 carcinomas were analyzed against paired normal tissue using the limma method^72^, genes significant (|logFC|>1 and FDR<0.01) in more than 2 carcinomas were included. 2. Enriched proteins from pan-cancer proteomic atlas^73^. 791 carcinoma samples collected from 15 tissues were included; each protein with logFC >1 and FDR<0.01 in more than 2 carcinomas was considered significant and included. 3. Carcinoma robust meta program markers: robust NMF was conducted in large-scale pan-cancer scRNA-seq across 18 tissues^74^, the characteristic genes for carcinoma meta-programs across more than two tissues were included. 4. Cell markers in the pan-cancer TME, nine main cell types in TME and 146 related subtypes from the latest pan-cancer scRNA-seq atlas, the top 10 genes in each subtype were included. We focused on genes that showed consistent evidence across genetic multi-omics analyses and that were also supported by these external resources, reasoning that such genes are strong candidates for shared carcinogenesis mechanisms and potential as biomarkers across carcinomas. Each gene providing more than 3 lines of evidence (including ≥1 line of genetic evidence) was considered a high-confidence genetic pleiotropic effector. Enrichment of these prioritized genes in known pathways or functional categories was examined by using Metascape^75^. We mainly report the top 20 clusters in pathway and process enrichment analysis.

### Phenome-wide Mendelian randomization analysis

Finally, we explored potential causal relationships between the latent cancer factors and a broad range of complex traits using phenome-wide mendelian randomization (PWMR) analyses. We analyzed 1,649 non-cancer phenotypes measured in the MVP cohort (451,206 individuals of European ancestry)^53^, spanning 22 phenotype categories. For each phenotype, we selected independent genome-wide significant SNPs for each exposure trait (P < 5×10^−8, LD r^2 < 0.001 within a 10 Mb window) as instrumental variables (IVs) using TwoSampleMRv0.6.2^76,77^. We excluded any IV that was palindromic with intermediate allele frequency or not present in the outcome GWAS^76^, that had low instrument strength (F-statistic < 10)^78^, or that was identified as an outlier with pleiotropy^79^. Causal effect estimates were primarily obtained using the inverse-variance weighted method (for exposures with >1 IV) or the Wald ratio method (for single-IV exposures). We additionally conducted sensitivity analyses using the weighted median and weighted mode estimators (when sufficient IVs were available) to ensure robustness of the findings^77^. We applied a stringent Bonferroni correction (α = 0.05/ (1,649*5) ≈ 6.06E−6) to identify statistically significant causally associated traits. We further required that a trait show concordant effect directions with nominal significance across all MR sensitivity methods to be considered a credible causal association. The merged UK10K–1KG reference panel^45,46^ was used.

## Ethics

Ethical approval for this study was not required as our analyses were based on summary statistics from published GWAS or the data were publicly accessible.

## Funding

Joint Funds for the Innovation of Science and Technology, Fujian Province (Grant number: 2023Y9299, to Chenshen Huang).

## Data Availability

Data availability
Summary GWAS carcinoma data are available from: GWAS Catalogue ID: GCST010098 (breast cancer); GWAS Catalogue ID: GCST90274714 (prostate cancer); GWAS Catalogue ID: GCST004462 (ovarian cancer); GWAS Catalogue ID: GCST006464 (endometrial cancer); GWAS Catalogue ID: GCST004748 (lung cancer); GWAS Catalogue ID: GCST90320057 (kidney cancer) ; GWAS Catalogue ID: GCST90399736 (thyroid cancer) ; GWAS Catalogue ID: GCST003739 (esophageal cancer); The reference panel, which is the merged genotype data from UK10K and 1000 Genomes Project Phase 3, is available from the EGA under accession number EGAD00001000776. Summary statistics from the latent factor GWAS will be uploaded to Zenodo and released upon online publication of the article. The remaining data are available within the article, Supplementary Information, or Source Data files.

## Acknowledgements

We thank the Dr.Kun-Long Huang reviewed the manuscript We thank the participants and coordinating teams of the Million Veteran Program (MVP), UK Biobank, FinnGen, and collaborating cancer genetics consortia whose data and infrastructure made this work possible. This research used data from the MVP, Office of Research and Development, U.S. Department of Veterans Affairs; the contents do not represent the views of the U.S. Department of Veterans Affairs or the United States Government. We analyzed publicly available UK Biobank GWAS summary statistics produced by the Neale Lab (Round 2, 2018), which were generated using the UK Biobank Resource under Applications 18597 and 11898; we did not access individual-level UK Biobank data. We used FinnGen R12 GWAS summary statistics; FinnGen is funded by Business Finland and industry partners as listed by the consortium, with samples provided by Finnish biobanks coordinated by BBMRI.fi/FinBB. For cancer GWAS resources, we acknowledge the Breast Cancer Association Consortium (BCAC) and the Consortium of Investigators of Modifiers of BRCA1/2 (CIMBA); the International Lung Cancer Consortium (ILCCO); the Genetics and Epidemiology of Colorectal Cancer Consortium (GECCO), the Colorectal Transdisciplinary Study (CORECT), and the Colon Cancer Family Registry (CCFR); the Barrett’s and Esophageal Adenocarcinoma Consortium (BEACON); the Global Biobank Meta-analysis Initiative (GBMI) for thyroid cancer; the Endometrial Cancer Association Consortium (ECAC) and the Epidemiology of Endometrial Cancer Consortium (E2C2); and the PRACTICAL consortium for prostate cancer. Where applicable, full study and funder lists are provided in the originating publications.

The breast cancer genome-wide association analyses for BCAC and CIMBA were supported by Cancer Research UK (PPRPGM-Nov20\100002, C1287/A10118, C1287/A16563, C1287/A10710, C12292/A20861, C12292/A11174, C1281/A12014, C5047/A8384, C5047/A15007, C5047/A10692, C8197/A16565) and the Gray Foundation, The National Institutes of Health (CA128978, X01HG007492-the DRIVE consortium), the PERSPECTIVE project supported by the Government of Canada through Genome Canada and the Canadian Institutes of Health Research (grant GPH-129344) and the Ministère de l’Économie, Science et Innovation du Québec through Genome Québec and the PSRSIIRI-701 grant, the Quebec Breast Cancer Foundation, the European Community’s Seventh Framework Programme under grant agreement n° 223175 (HEALTH-F2-2009-223175) (COGS), the European Union’s Horizon 2020 Research and Innovation Programme (634935 and 633784), the Post-Cancer GWAS initiative (U19 CA148537, CA148065 and CA148112 - the GAME-ON initiative), the Department of Defence (W81XWH-10-1-0341), the Canadian Institutes of Health Research (CIHR) for the CIHR Team in Familial Risks of Breast Cancer (CRN-87521), the Komen Foundation for the Cure, the Breast Cancer Research Foundation and the Ovarian Cancer Research Fund. All studies and funders are listed in Zhang H et al (Nat Genet, 2020).

## Author contributions

H.Y.F. designed the analysis; H.Y.F. performed the data analysis and wrote the original draft; H.Y.F. and H.C. reviewed and edited the manuscript; H.C. acquired funding.

## Declaration of Interests

All authors declare no competing interests.

## Data availability

Summary GWAS carcinoma data are available from: GWAS Catalogue ID: GCST010098 (breast cancer); GWAS Catalogue ID: GCST90274714 (prostate cancer); GWAS Catalogue ID: GCST004462 (ovarian cancer); GWAS Catalogue ID: GCST006464 (endometrial cancer); GWAS Catalogue ID: GCST004748 (lung cancer); GWAS Catalogue ID: GCST90320057 (kidney cancer) ; GWAS Catalogue ID: GCST90399736 (thyroid cancer) ; GWAS Catalogue ID: GCST003739 (esophageal cancer); The reference panel, which is the merged genotype data from UK10K and 1000 Genomes Project Phase 3, is available from the EGA under accession number EGAD00001000776. Summary statistics from the latent factor GWAS will be uploaded to Zenodo and released upon online publication of the article. The remaining data are available within the article, Supplementary Information, or Source Data files.

## Code availability

Custom analysis scripts are hosted on GitHub (https://github.com/1667857557/Cross-carcinoma-genetic-pleiotropic-effects).

## Notes

### Competing Interest Statement

The authors have declared no competing interest.

### Funding Statement

This study did not receive any funding

### Summary of Updates

Funding details have been added, and minor revisions were made to the figures and text to improve layout and clarity.

