## Supplement Figure 1 to 9 for "Multivariate GWAS reveals shared genetic etiology and pleiotropic loci across carcinomas"

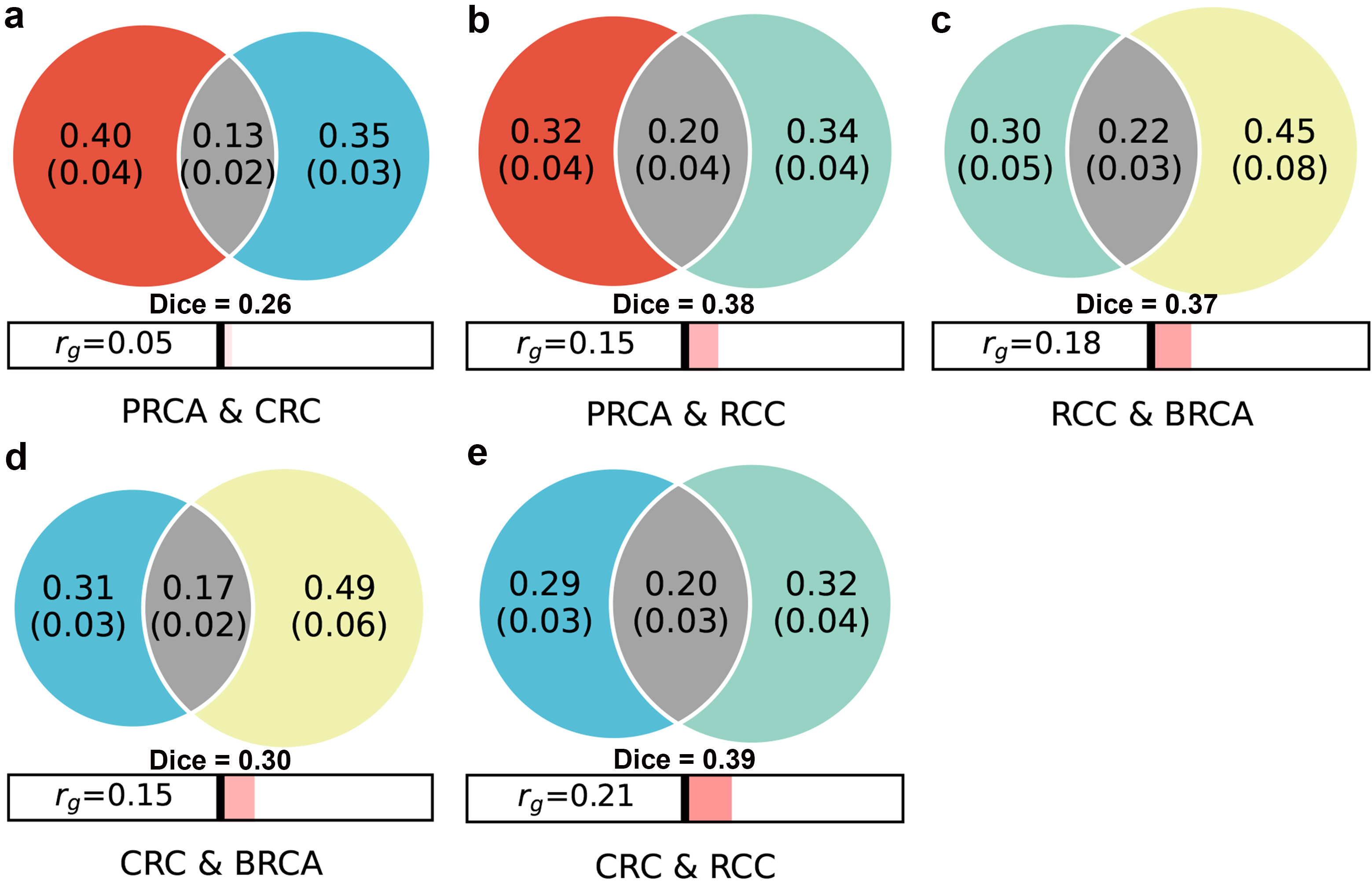

**Fig S1 |** **Polygenic overlap between carcinoma pairs estimated by bivariate MiXeR.**

*(a) PRCA–CRC; (b) PRCA–RCC; (c) RCC–BRCA; (d) CRC–BRCA; (e) CRC–RCC.* Venn diagrams show MiXeR-estimated numbers of trait-influencing (non-null) variants unique to each cancer and shared between them; point estimates are shown with s.e. in parentheses. Horizontal bars below each panel summarize the genome-wide genetic correlation (𝑟𝑔). The Dice coefficient quantifies the relative proportion of shared elements between two sets.

Abbreviations: PRCA, prostate cancer; CRC, colorectal cancer; RCC, renal cell cancer; BRCA, breast cancer.
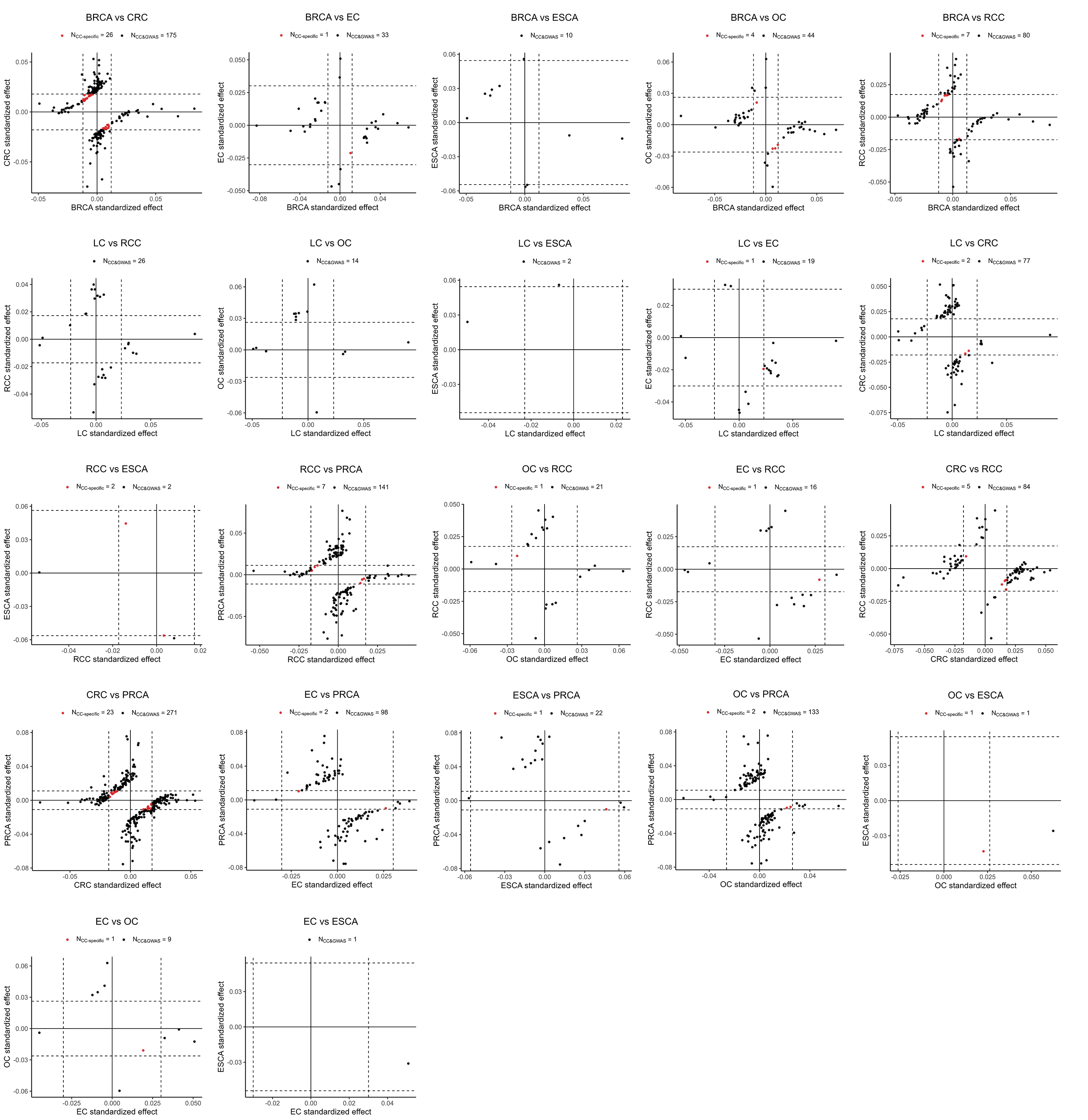

**Fig S2 | Pairwise CC-GWAS contrasts across other carcinomas.** Scatterplots show standardized per-allele effects at LD-independent lead loci from each CC-GWAS contrast. For each panel, the x- and y-axes give the standardized effects from the corresponding case–control GWAS of the first- and second-named cancers, respectively; solid lines mark zero and dashed lines indicate the study-specific significance thresholds used in the case–control scans. Red points denote case–case–specific loci (genome-wide significant in CC-GWAS but not significant in either case–control GWAS for the contrasted cancers), whereas black points denote loci detected by both CC-GWAS and at least one underlying case–control GWAS. Numbers in each panel report counts of case–case–specific and shared loci. Quadrants classify directionality: upper-right and lower-left indicate concordant risk directions across cancers; upper-left and lower-right indicate opposite risk directions, consistent with differential genetic effects. Effects were harmonized to the same effect allele across studies.

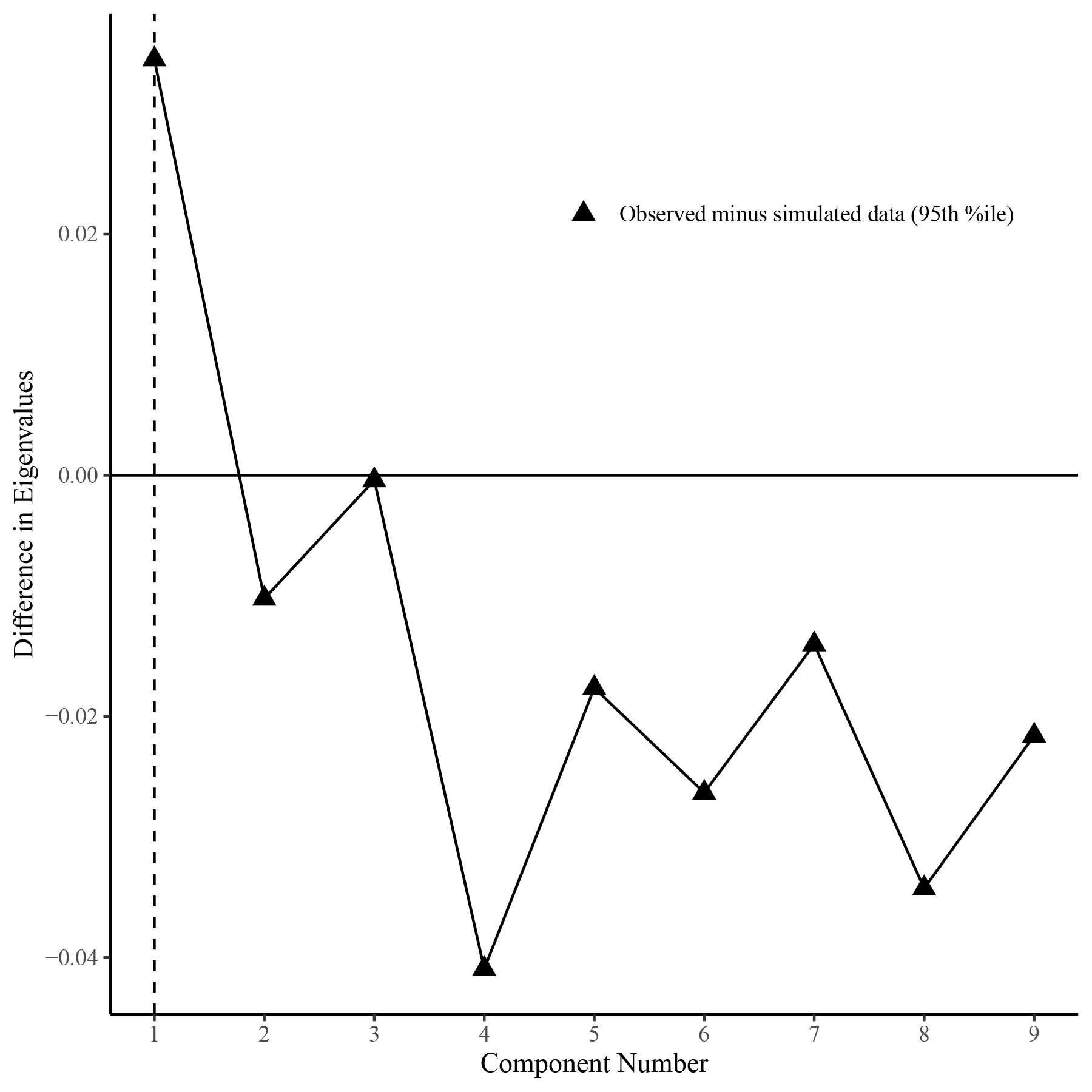

**Fig S3 | Parallel analysis of LDSC genetic covariance to select factor dimensionality.** Shown is the difference between the observed eigenvalues of the LDSC genetic covariance matrix S and the 95th percentile of eigenvalues from 100 parametric bootstrap datasets simulated from the sampling covariance V (triangles). The horizontal line marks zero; positive values indicate components exceeding chance expectation. The dashed vertical line denotes the retained dimensionality (here k=1), as the first component was the largest. Analysis was performed with genomicSEM’s paLDSC using ldsc.covstruct (nine traits).

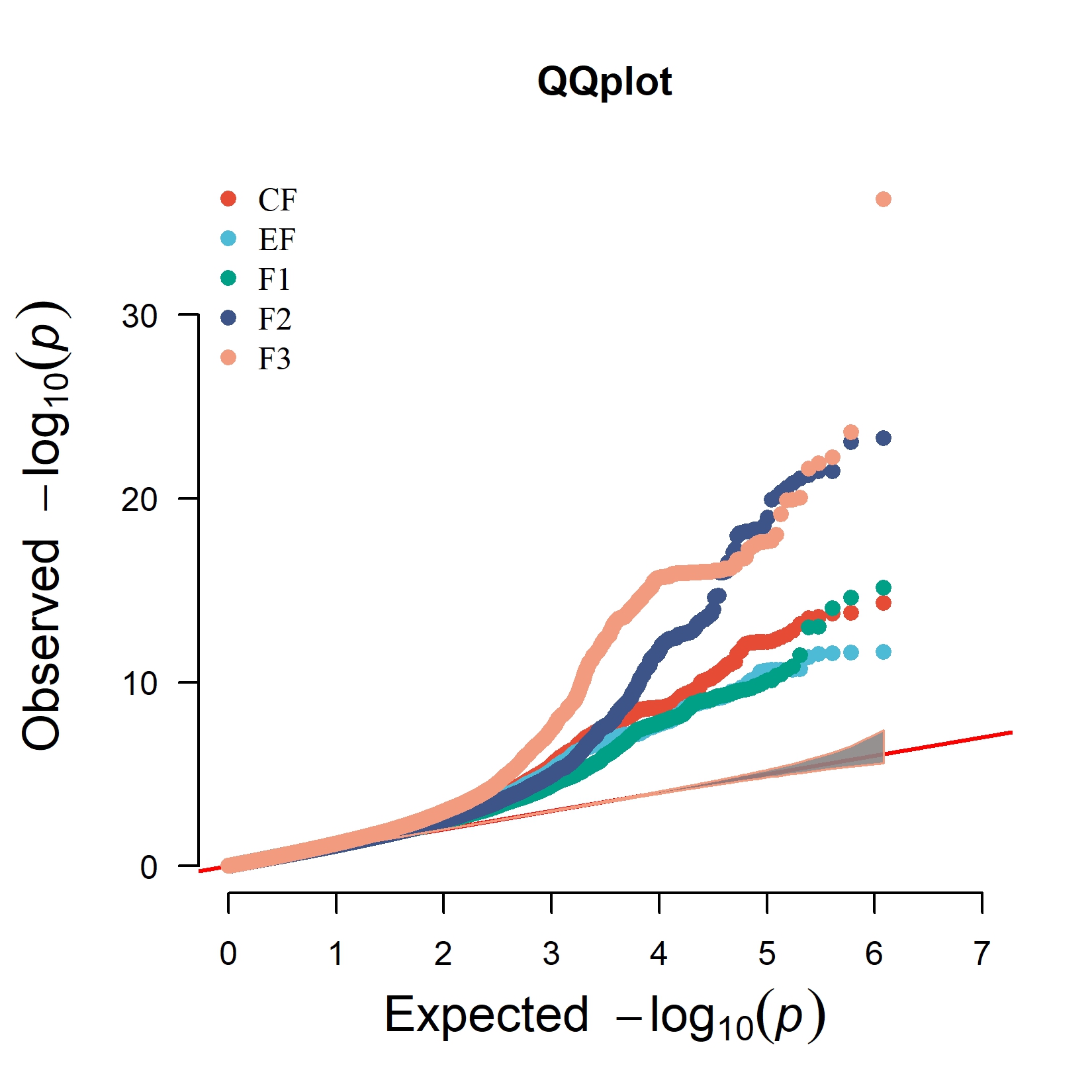

**Fig S4 | Quantile–quantile plots for latent-factor GWAS.** Observed versus expected −log10(p) for genome-wide association tests of five latent factors (CF, EF, F1, F2, F3; colours as indicated). The red diagonal denotes the null expectation; the grey envelope marks the 95% sampling bounds under the null. Curvature above the diagonal indicates an excess of small ppp values consistent with polygenicity and lead associations, whereas alignment with the diagonal at lower quantiles indicates well-calibrated test statistics. Full GWAS details (sample sizes, covariates and quality control) are provided in the Methods.

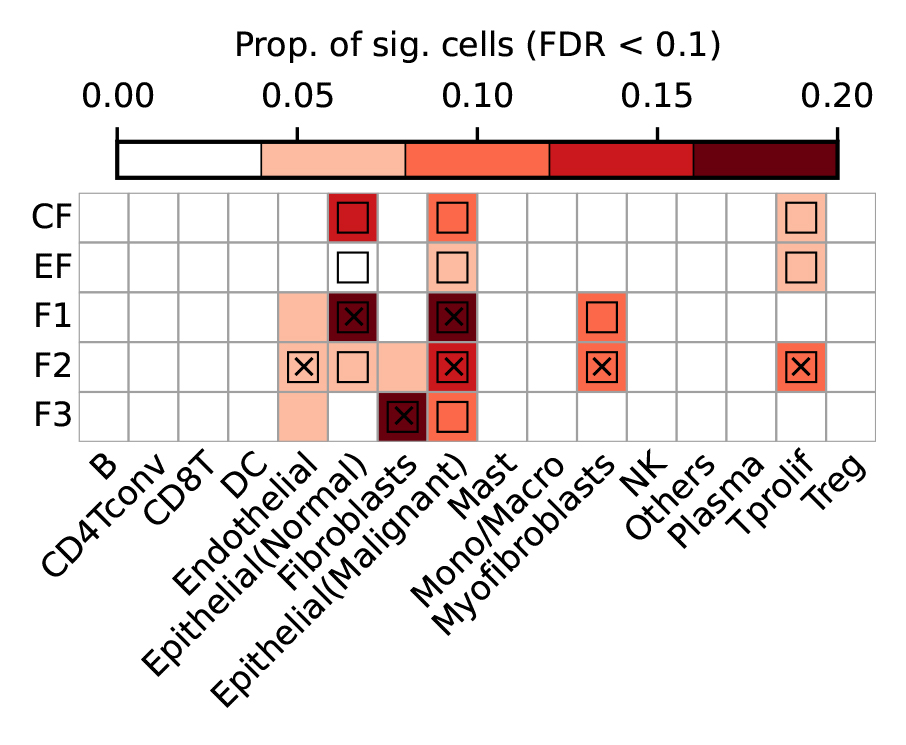

**Fig S5 | Heatmap plots of scDRS analysis in latent-factor GWAS**

Heatmap colors for each cell type-disease pair denote the proportion of significantly associated cells. Squares denote significant cell type-disease associations (FDR <0.05 across all pairs of cell types and carcinomas. Cross symbols denote significant heterogeneity in association with disease across individual cells within a given cell type.

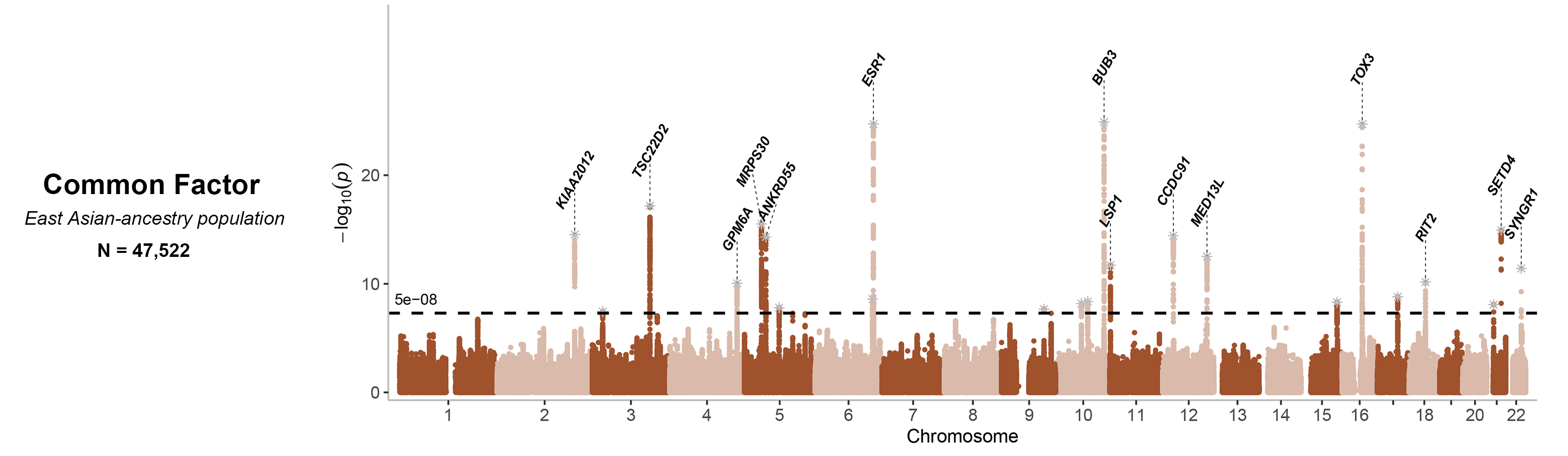

**Fig S6 | Manhattan plots of GenomicSEM East Asian-ancestry population common factor GWAS.**
Each panel shows –log10(P) for SNP associations across autosomes (chr 1–22); the dashed line marks genome-wide significance (P = 5 × 10⁻⁸). Alternating background shading distinguishes chromosomes. Lead variants at genome-wide significant loci are annotated by nearest gene. Panel titles report the effective sample size (N) for common factor GWAS.

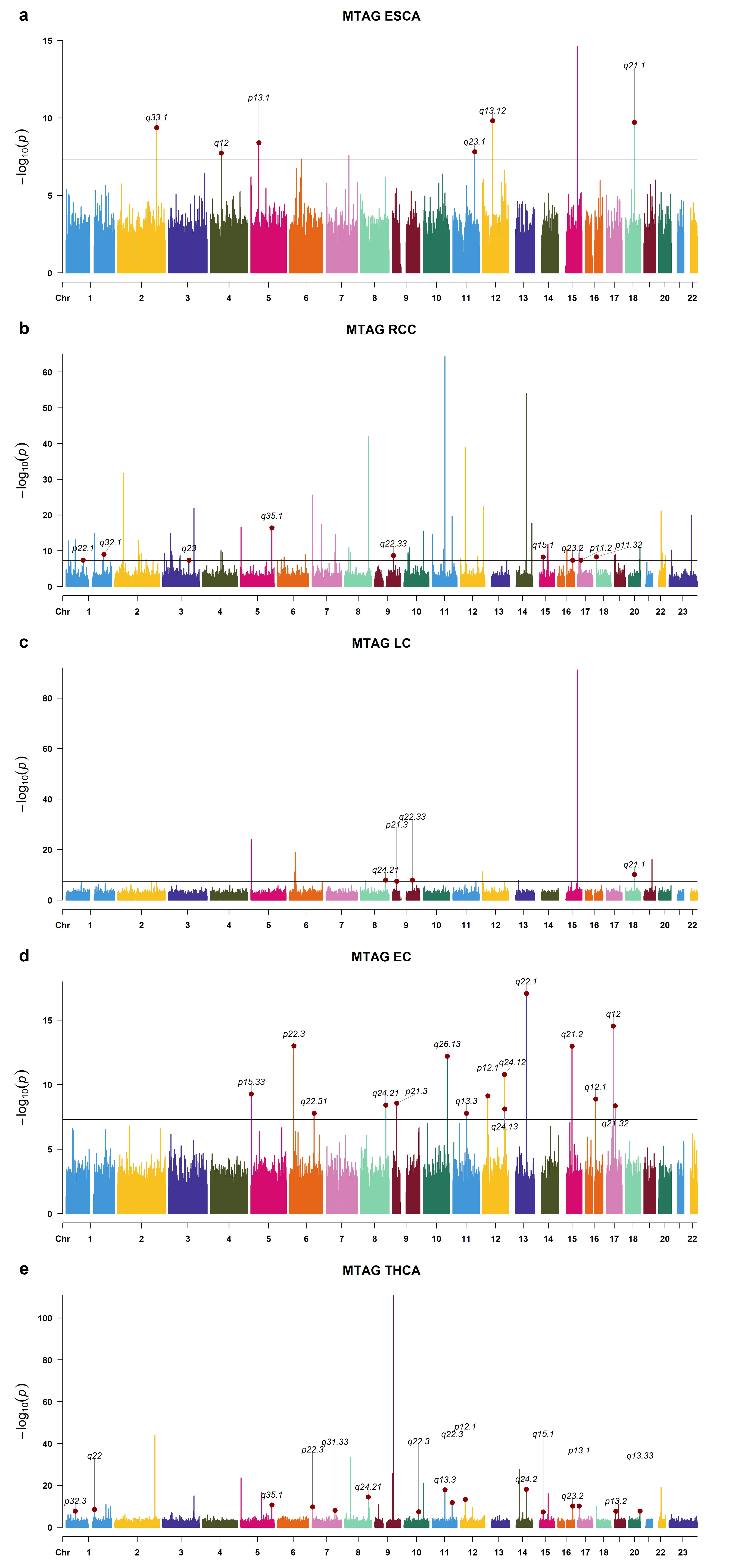

**Fig. S7| Manhattan plots of multi-trait multivariate GWAS.**

a–e Manhattan plots for MTAG results of each novel loci identified cancer GWAS. The X-axis represented the chromosomal position, and the Y-axis showed the negative log10-transformed P values for each SNP. Cytoband annotations for the newly identified genomic loci are shown in gray. Genome-wide significance threshold of P = 5 × 10−8 (black dotted line). Darkred dots indicated an independent genome-wide significant association with the smallest P value (Top lead SNP). All statistical tests are two-sided. MTAG multi-trait analysis of GWAS.

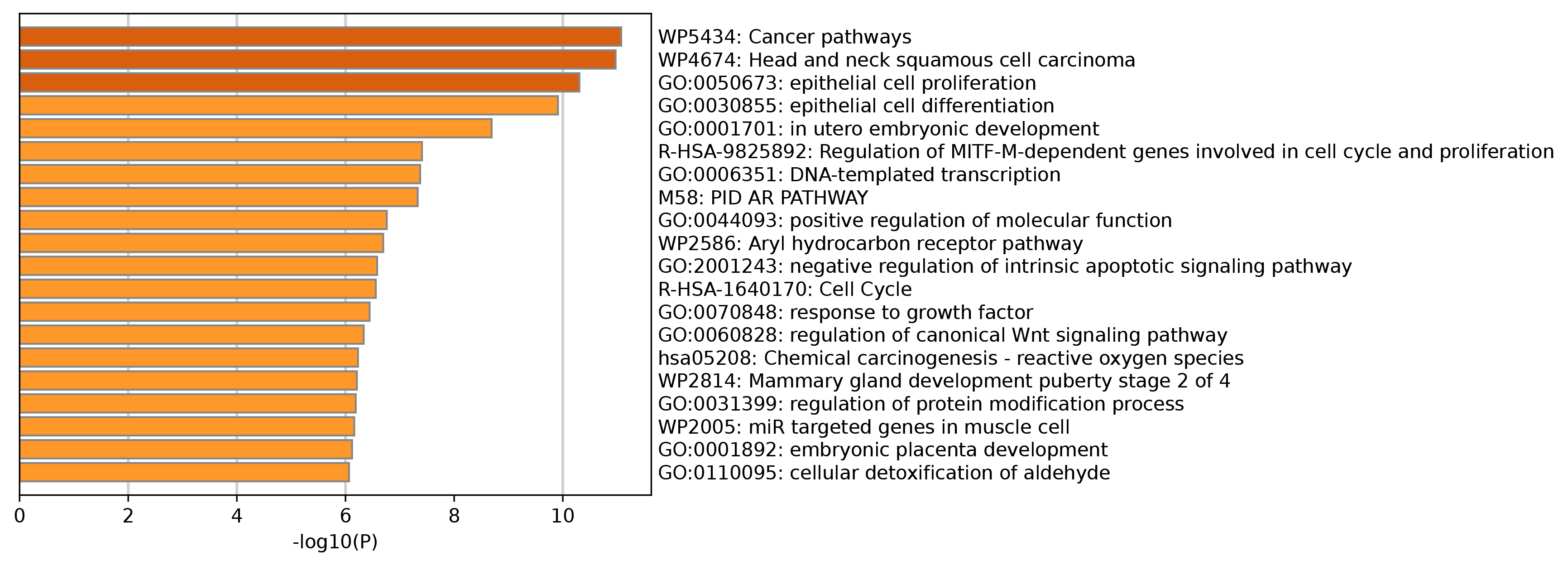

**Fig S8 | Pathway enrichment of 167 high-confidence pleiotropic effector genes.** Horizontal bars show the top enriched Gene Ontology biological processes and curated pathway terms (GO, Reactome, KEGG, WikiPathways, MSigDB), ranked by −log10(P). Enrichment was tested with a hypergeometric model against all protein-coding genes and P values were Benjamini–Hochberg FDR–adjusted; the vertical line marks the 5% FDR threshold.

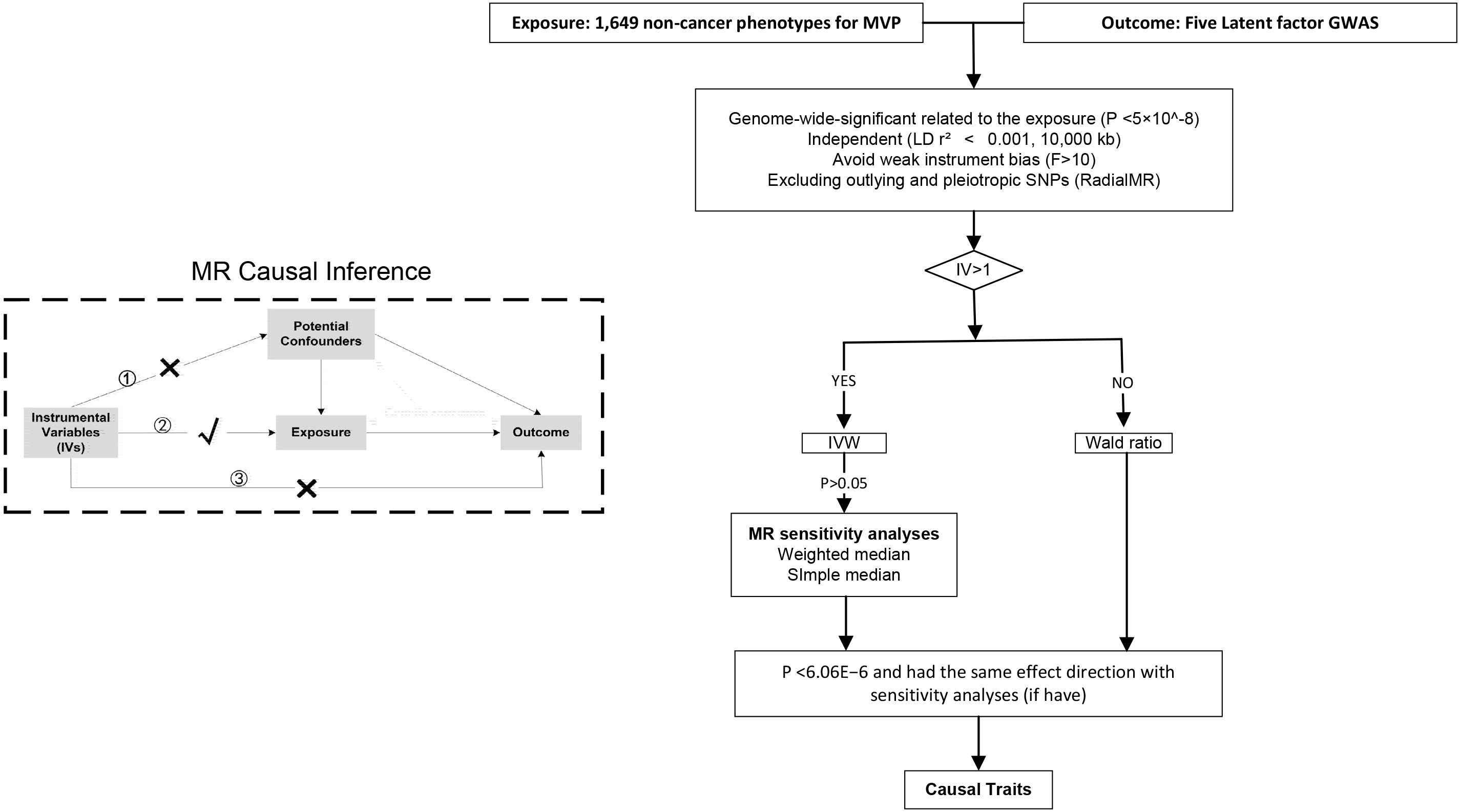

**Fig S9 |** **Phenome-wide Mendelian randomization analysis.** When exposure and outcome are used for causal estimates in MR inference, three assumptions must be satisfied. ① Relevance assumption: the genetic variations are highly related to the exposure, ② Independence assumption: the genetic variants are not associated with any putative confounder of the association between exposure and outcome, and ③ Exclusion restriction: the variants do not alter the outcome independently of exposure.
